# Improving estimates of social contact patterns for the airborne transmission of respiratory pathogens

**DOI:** 10.1101/2022.02.06.22270386

**Authors:** Nicky McCreesh, Mbali Mohlamonyane, Anita Edwards, Stephen Olivier, Keabetswe Dikgale, Njabulo Dayi, Dickman Gareta, Robin Wood, Alison D Grant, Richard G White, Keren Middelkoop

## Abstract

**Background:** Data on social contact patterns are widely used to parameterise age-mixing matrices in mathematical models of infectious diseases designed to help understand transmission patterns or estimate intervention impacts. Despite this, little attention is given to how social contact data are collected and analysed, or how the types of contact most relevant for transmission may vary between different infections. In particular, the majority of studies focus on close contacts only – people spoken to face-to-face. This may be appropriate for infections spread primarily by droplet transmission, but it neglects the larger numbers of ‘shared air’ casual contacts who may be at risk from airborne transmission of pathogens such as *Mycobacterium tuberculosis*, measles, and SARS-CoV-2.

**Methods:** We conducted social contact surveys in communities in two provinces of South Africa in 2019 (KwaZulu-Natal and Western Cape). In line with most studies, we collected data on people spoken to (close contacts). We also collected data on places visited and people present, allowing casual contact patterns to be estimated. Using these data, we estimated age mixing patterns relevant for i) droplet and ii) non-saturating airborne transmission. We also estimated a third category of pattern relevant for the transmission of iii) *Mycobacterium tuberculosis* (*Mtb*), an airborne infection where saturation of household contacts plays an important role in transmission dynamics.

**Results:** Estimated contact patterns by age did not vary greatly between the three transmission routes/infections, in either setting. In both communities, relative to other adult age groups, overall contact intensities were lower in 50+ year olds when considering contact relevant for non-saturating airborne transmission or the transmission of *Mycobacterium tuberculosis* than when considering contact relevant for droplet transmission.

**Conclusions:** Our findings provide some reassurance that the widespread use of close contact data to parameterise age-mixing matrices for transmission models of airborne infections may not be resulting in major inaccuracies. The contribution of older age groups to transmission may be over-estimated, however. There is a need for future social contact surveys to collect data on casual contacts, to investigate whether our findings can be generalised to a wider range of settings, and to improve model predictions for infections with substantial airborne transmission.

## Introduction

Mathematical models of infectious disease transmission are widely used to help inform infectious disease policy, estimate the potential impact of interventions, and to provide insight into disease dynamics and natural history. Many models incorporate patterns of mixing between different sections of the population, most commonly between different age groups. Simulated mixing patterns can have a large effect on model dynamics(1), and it is therefore important that realistic mixing patterns are simulated. Mixing patterns are frequently informed by social contact data: empirical data collected from respondents on the people that they had contact with over a set period of time(2).

The majority of social contact data collection has focused on ‘close’ contacts, with most studies using a definition of contacts that required a two way face-to-face conversation of at least three words, close proximity (e.g. within 2m), and/or physical contact(2). It is plausible that these types of contact approximate reasonably well the types of contact that are relevant for the transmission of infections that are transmitted primarily through direct contact and droplets. For obligate, preferential, or opportunistic airborne infections such as measles, *Mtb*, and SAR-CoV-2, however, it is likely that this definition excludes many potentially effective contacts. This is because transmission of airborne infections can occur between anybody ‘sharing air’ in inadequately ventilated indoor spaces, regardless of whether conversation occurs, and over distances of more than 2m(3). For airborne infections, estimates of ‘casual contact’ time may therefore be more appropriate, calculated as the time spent in indoor locations multiplied by the number of other people present.

Tuberculosis also differs from the majority of respiratory infections in the long durations of time for which people are potentially infectious – with an estimated 9-36 months between disease development and diagnosis/notification across 11 high burden countries(4). This means that transmission to repeated contacts can partially saturate (even allowing for reinfection), making the relationship between contact time and infection risk non-linear(5). This effect is most pronounced for contact between household members(5). Household membership and repeated contacts are rarely explicitly simulated in mathematical models, and therefore the effects of contact saturation need to be incorporated into the mixing matrices used to parameterise the models.

In this paper, we describe methods for estimating age mixing patterns relevant for non-saturating airborne transmission and *Mtb*, using a novel weighted approach to incorporate the effects of household contact saturation into our estimates for *Mtb*. We generate estimates of age mixing using data on close and casual contacts from two communities in South Africa, and compare the estimated mixing patterns with those typically used in mathematical modelling studies – generated using close contact numbers, and more suitable for the droplet transmission.

## Methods

Social contact data were collected in two study communities in South Africa: one in KwaZulu-Natal Province, and one in Western Cape Province. Both communities have high rates of unemployment, and high prevalences of HIV and incidences of tuberculosis compared to the provinces as a whole. The study community in KwaZulu-Natal consisted of a population of 46,000, living in the predominantly rural and peri-urban areas in the catchment areas of two primary care clinics, and within a demographic surveillance area (DSA). The study community in Cape Town was a peri-urban community of 27,000 people, and was an established research site with biennial censuses.

### Data collection

The KwaZulu-Natal data were collected between March and December 2019. 3093 adults (aged 18+ years) were sampled at random, stratified by residential area (∼350 households per area) and with probability proportional to the number of eligible people in each area, based on the most recent DSA census conducted prior to area entry. Up to three attempts were made to contact sampled individuals.

The Western Cape data were collected in May to October 2019. In total, 1530 adults (aged 15+ years) were selected using age- and sex-stratified random sampling, based on a census conducted in the study population in February and March 2019. Up to five attempts were made to contact selected individuals. Each attempt was made on different days of the week (including at weekends), at different times of day to maximise the chance of making contact.

For both surveys, interviews were conducted face-to-face at the respondents’ homes, using interview administered questionnaires on tablet computers. Interviews were conducted in isiZulu in KwaZulu Natal, and in English or isiXhosa in Western Cape. Respondents were asked about their movements on a randomly assigned day in the preceding week in KwaZulu-Natal, and on the day before the interview in Western Cape. To allow casual contact time (defined as time spent ‘sharing air’ indoors or on transport) to be estimated, respondents were asked to list the places they had visited (including their own home) and transport they had used. For each location, questions asked included:

- What type of location was it?
- How long did you spend there?
- How many people were there, halfway through the time you were there?

Respondents were also asked about their close contacts, defined as people that the respondent had a face-to-face conversation with. Respondents were first asked to make a list of all their contacts, with help from the interviewer. Respondents were then asked questions about a random 10 contacts, or all of their contacts if they reported fewer than 10. Questions included:

- Is this contact a member of your household?
- How old do you think they are?
- How much time did you spend with them in total?

Respondents’ basic demographic information were also collected. For the KwaZulu-Natal community, data on household size and residency (urban, peri-urban, rural) were obtained from the most recent DSA census. All other data were collected directly from the respondents.

### Ethics

Ethical approval for the data collection in KwaZulu-Natal was granted by the Biomedical Research Ethics Committee (BREC) of the University of KwaZulu-Natal (BE662/17) and the London School of Hygiene & Tropical Medicine Observational / Interventions Research Ethics Committee (14640). Ethical approval for the data collection in Western Cape was granted by the Human Research Ethics Committee at the University of Cape Town (HREC/REF: 008/2018) and the London School of Hygiene & Tropical Medicine Observational / Interventions Research Ethics Committee (14520). Informed consent was obtained from all participants.

### Data analysis

Close contact numbers and times were estimated using data on people with whom the respondents reported having a face-to-face conversation. 95% plausible intervals for the age mixing matrices were generated using bootstrapping. Household and non-household age-mixing matrices were generated from sampled contacts who were and were not reported to be members of the respondents’ own households respectively.

Casual contact time in a location was estimated as the duration of time the respondent reported spending there, multiplied by the reported number of people present. Central estimates for casual contact time age-mixing matrices were generated using the method outlined in McCreesh *et al*(6). In brief, as data were collected on numbers of total people and children present in indoor locations only, and not the ages of adults, the age distribution of adult casual contacts needed to be estimated. We therefore assumed that the age distribution of adult contacts in each location type matched the age distribution of respondents who reported visiting location of that time, weighted by the duration of time they reported spending in that location time, and weighted to the sampled population age and sex distribution. Household and non-household contact matrices were generated using data on contact time in respondents’ own homes and all other locations respectively. Plausible ranges were generated using bootstrapping.

The age mixing matrices were adjusted to be symmetrical, using the study community age structures. Data on adult contact numbers and time with children were used to estimate child contact numbers and time with adults. To allow comparison between the two study communities, the lowest respondent age group was set at 15-19 years for both surveys. As 15-17 year olds were not interviewed in KwaZulu-Natal, we assumed that contact patterns in 18-19 years olds were representative of contact patterns in all 15-19 year olds.

Further details of the analysis methods are given in the appendix.

### Generating age-mixing matrices for droplet and non-saturating airborne transmission, and *Mycobacteria tuberculosis*

Figure 1 summarises the data used to generate the age mixing matrices for droplet and non-saturating airborne transmission, and *Mtb*.

**Figure 1.**
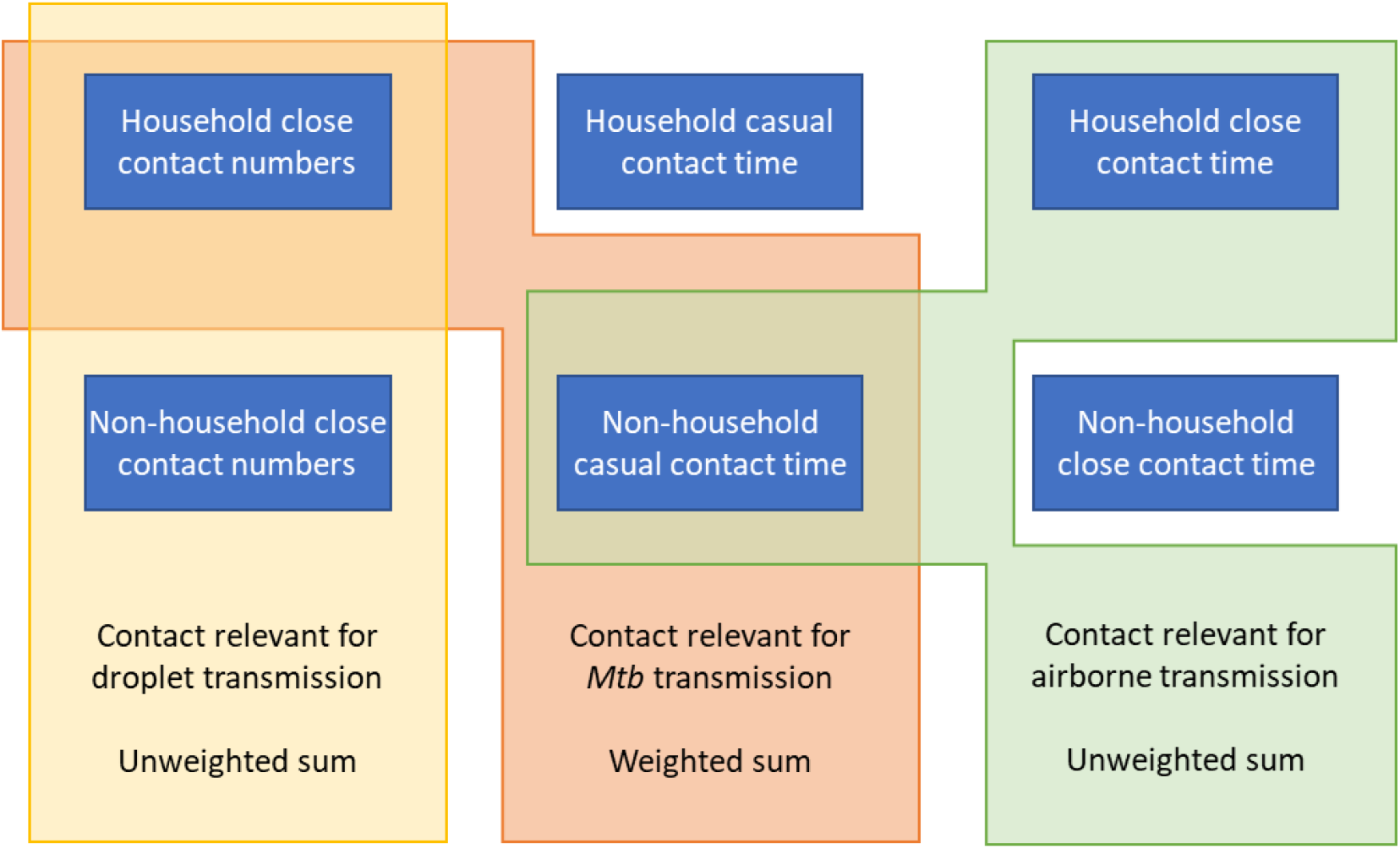
Summary of data used to estimate age-mixing matrices. Diagram showing how age-mixing matrices relevant for the transmission of droplet infections, airborne infections, and *Mycobacterium tuberculosis* were estimated using empirical data on close contact numbers, close contact time, and casual contact time.

Age-mixing matrices relevant for droplet transmission were set equal to age-mixing matrices calculated using close contact numbers.

Age-mixing matrices relevant for non-saturating airborne transmission were set equal to the unweighted sum of the household close contact time matrices and the non-household casual contact time matrices. Close contact time was used for household estimates as 1) it was considered plausible that the majority of contact between household members is likely to meet the definition of ‘close contact’, and 2) it allowed the age structures of households to be more accurately reflected in the age-mixing matrices.

Age-mixing matrices relevant for *Mtb* were set equal to the sum of the household close contact number matrices and the non-household casual contact time matrices, weighted to reflect empirical estimates of the proportion of tuberculosis that results from household transmission. For each pair of bootstrapped household and non-household matrices, a proportion of ‘contact’ that should occur in households was sampled from a uniform distribution between 8-16% (with 12% used for the central estimate)(5). A weighted average was then generated, given the desired proportion of overall ‘contact’ by adults occurring in households.

To allow direct comparisons to be made between the different age mixing matrices, the matrices for non-saturating airborne transmission and *Mtb* were adjusted to give the same mean contact intensity between adults as the matrices for droplet transmission.

Further details of the analysis methods are given in the appendix.

## Results

### Recruitment

Of the 3093 people sampled in KwaZulu-Natal, 1723 (56%) were successfully contacted, 299 (10%) were dead or reported to have out-migrated, and 1071 (35%) could not be contacted. Of those successfully contacted, 1704 (99%) completed an interview.

Of the 1530 people sampled in Western Cape, 1214 (93%) were successfully contacted, 117 (8%) had moved or died, 193 (13%) had had incorrect information listed in the census, and 6 were uncontactable. Of the 1214 people contacted, 77 (6%) refused to be interviewed and 14 were ineligible (due to disability or lack of English and isiXhosa). Of 1123 people interviewed, technical issues meant that data from 8 interviews were lost, leaving 1115 (92%) completed interviews.

Table 1 shows the characteristics of the respondents and the target population. For both populations, the recruited sample was a reasonable match to the target population in terms of sex, age, and residence type (urban, peri-urban, or rural). Unemployment was high in both populations, with only 23% of respondents in KwaZulu-Natal and 55% in Western Cape reporting full-time, part-time, or casual employment. Household sizes were large in KwaZulu-Natal, with 48% living in a household of eight or more, and smaller in Western Cape, with 79% living in a household of four or fewer.

**Table 1.** Respondent characteristics. Target population refers to people in the populations aged 15+ years. *In KwaZulu-Natal, urban is defined as KwaMsane Municipality, peri-urban as other areas with a population density over 400/km^2^, and rural as areas with a population density under 400/km^2^

|  |  | KwaZulu Natal |  | Western<br>Cape |  |
| --- | --- | --- | --- | --- | --- |
|  |  | Sample | Target<br>population | Sample | Target<br>population |
| **Sex** | Male | 751 (44%) | 41% | 553 (50%) | 52% |
|  | Female | 953 (56%) | 59% | 562 (50%) | 48% |
| **Age** | 15-17 | 0 | 9.1% | 56 (5%) | 4.5% |
|  | 18-19 | 118 (6.9%) | 5.6% | 84 (7.5%) | 4.5% |
|  | 20-29 | 495 (29%) | 26% | 412 (37%) | 33% |
|  | 30-39 | 308 (18%) | 21% | 358 (32%) | 37% |
|  | 40-49 | 227 (13%) | 13% | 142 (13%) | 15% |
|  | 50+ | 556 (33%) | 25% | 63 (5.7%) | 6.5% |
| **Residence\*** | Rural | 867 (51%) | 59% | 0 | 0% |
|  | Peri-Urban | 716 (42%) | 33% | 1115 (100%) | 100% |
|  | Urban | 121 (7.1%) | 8% | 0 | 0% |
| **Monthly household income** | Less than R1000 | 416 (24%) |  | 111 (10%) |  |
|  | R1000 - R2500 | 785 (46%) |  | 261 (23%) |  |
|  | R2500 - R5000 | 302 (18%) |  | 374 (34%) |  |
|  | R5000 - R10000 | 125 (7.3%) |  | 179 (16%) |  |
|  | More than R10000 | 65 (3.8%) |  | 61 (5.5%) |  |
|  | Unknown/missing | 11 (0.65%) |  | 129 (12%) |  |
| **Employment** | Full-time | 329 (19%) |  | 403 (36%) |  |
|  | Part-time/Casual | 68 (4%) |  | 213 (19%) |  |
|  | None | 1299 (76%) |  | 492 (44%) |  |
|  | Missing | 8 (0.5%) |  | 7 (0.6%) |  |
| **Household size** | 1 | 115 (6.7%) | 4.1% | 203 (18%) | 19% |
|  | 2-4 | 287 (17%) | 26% | 683 (61%) | 66% |
|  | 5-7 | 488 (29%) | 33% | 195 (17%) | 13% |
|  | 8-10 | 375 (22%) | 20% | 26 (2.3%) | 1.6% |
|  | 11+ | 439 (26%) | 17% | 8 (0.72%) | 0.4% |
| **Day reported** | Monday | 239 (14%) |  | 203 (18%) |  |
|  | Tuesday | 242 (14%) |  | 202 (18%) |  |
|  | Wednesday | 239 (14%) |  | 187 (17%) |  |
|  | Thursday | 251 (15%) |  | 138 (12%) |  |
|  | Friday | 261 (15%) |  | 80 (7.2%) |  |
|  | Saturday | 245 (14%) |  | 98 (8.8%) |  |
|  | Sunday | 227 (13%) |  | 207 (19%) |  |
| **Total** |  | 1704 | 33,288 | 1115 | 20,633 |

### Contact numbers and time

Figure 2 and Appendix tables S1-S6 show household and non-household close contact numbers and time and casual contact time in KwaZulu-Natal and Western Cape, by sex, age, and household size. Overall close contact numbers and time, and casual contact time, were significantly higher for women than for men in both communities, however the differences were generally not large. There was a tendency for casual contact time to decrease slightly with age in both communities, and close contact numbers and time were substantially higher in 15-19 year olds than in older age groups in Western Cape. Close contact numbers and time, and casual contact time, increased with increasing household size in both settings, driven by increases in contact with household members. Differences by household size were larger in Western Cape than in KwaZulu-Natal, most likely reflecting the fact that household sizes were self-reported in Western Cape, but obtained through linking to community census data in KwaZulu-Natal. Contact between household members made up a higher proportion of total contact in KwaZulu-Natal than in Western Cape, for all types of contact.

**Figure 2.**
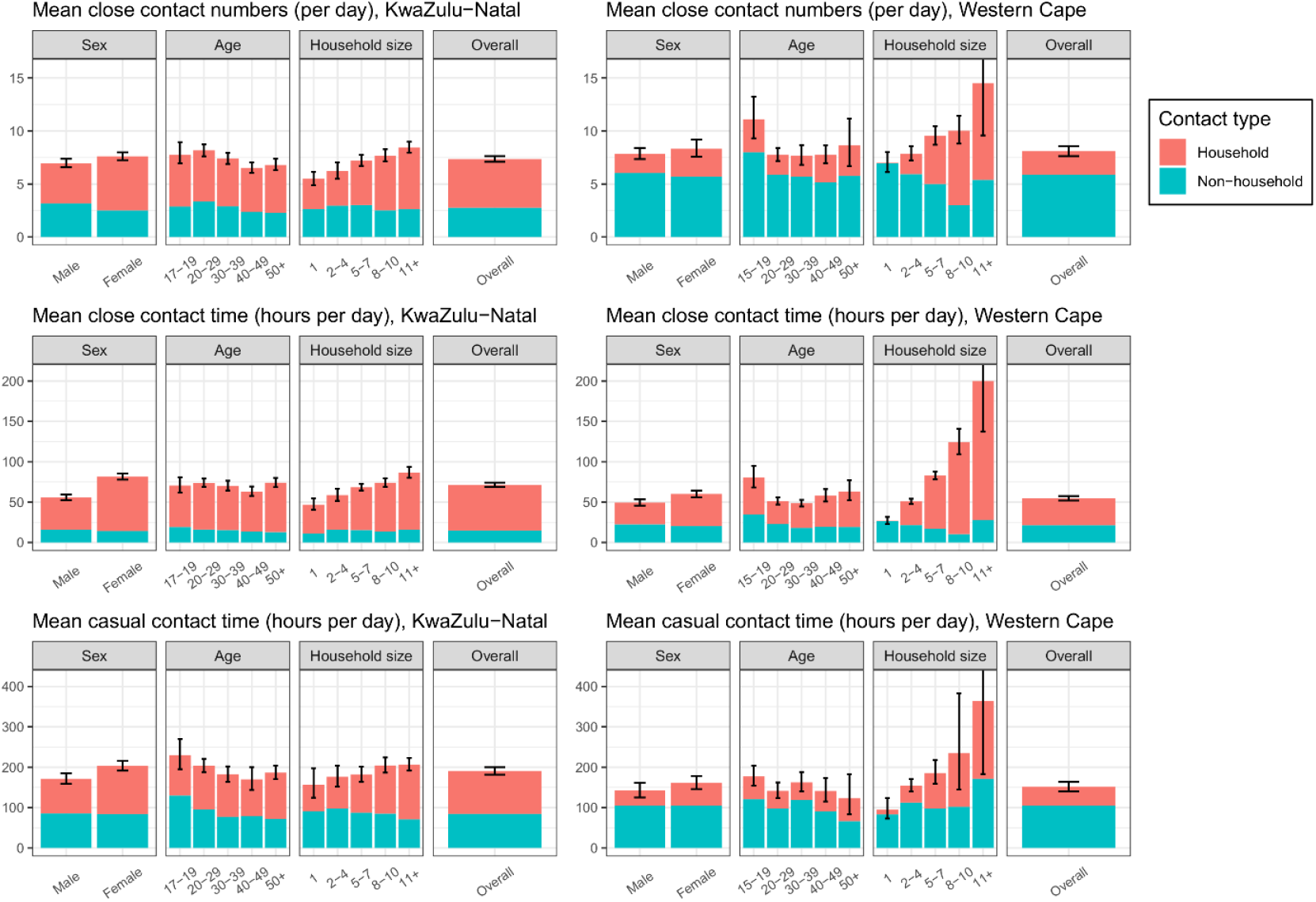
Household and non-household close contact numbers, close contact time, and casual contact time in KwaZulu-Natal and Western Cape, by sex, age and household size. Error bars show 95% confidence intervals for total contact numbers or time.

### Age mixing

Figure 3 and 4 show estimated age mixing matrices for droplet transmission, non-saturating airborne transmission, and *Mtb* transmission, for KwaZulu-Natal and Western Cape respectively.

**Figure 3.**
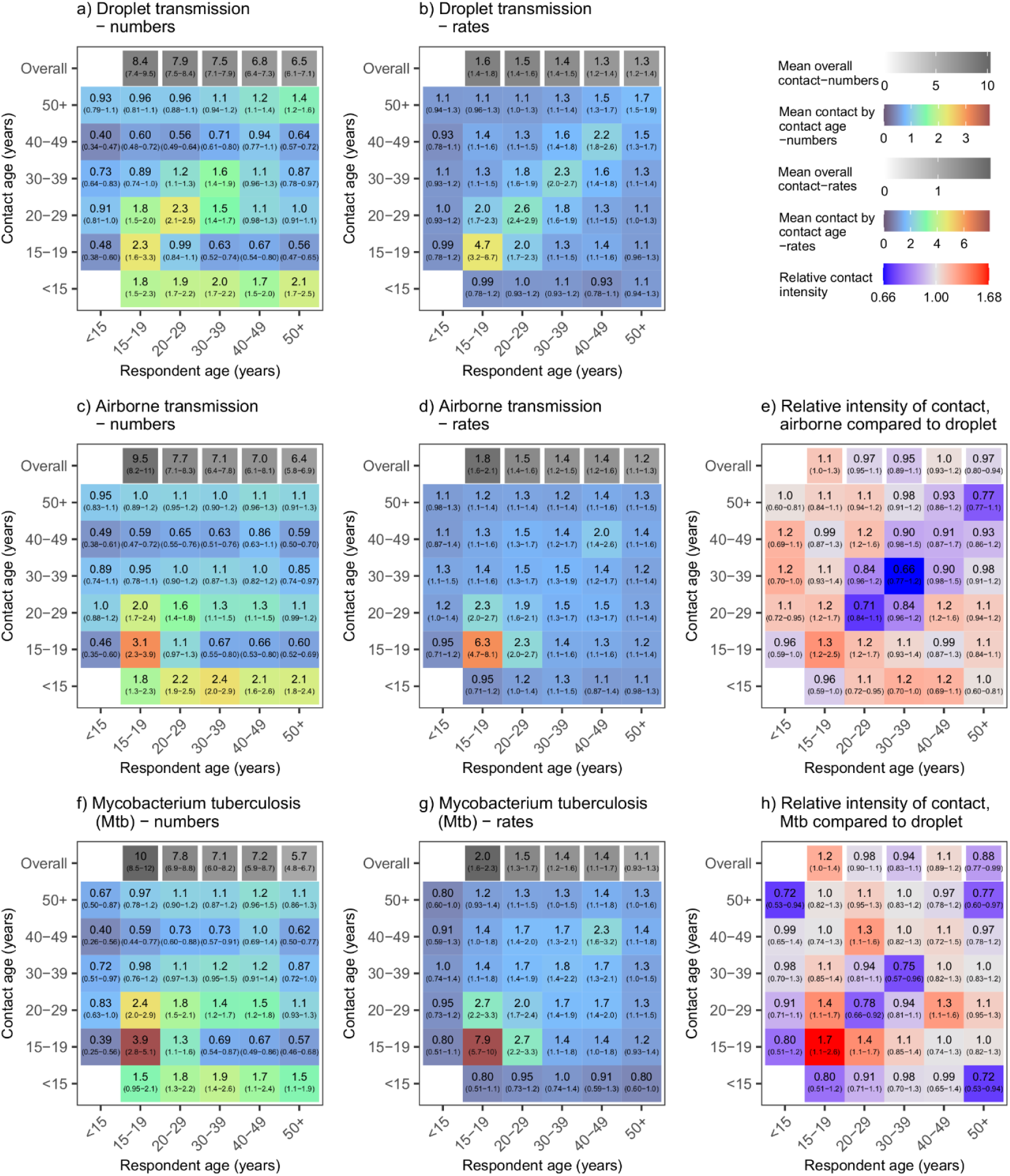
Age mixing matrices relevant for droplet transmission, non-saturating airborne transmission, and *Mycobacterium tuberculosis* transmission, for KwaZulu-Natal. Graphs a, c, and f show absolute contact intensities between respondents and contacts in each age group. Graphs b, d, and g show intensities of contact between each member of each age group. Graphs e and h show intensities for airborne infections and *Mtb* compared to intensities for droplet infections respectively. Numbers shown in graph a are the mean number of contacts respondents in each age group have with contacts in each age group per day. Numbers shown in graph b are the rate of contact between each individual in the population per day, expressed as rates ×10^5^. ‘Numbers’ and ‘rates’ in graphs c, d, f, and g are standardised so that the mean overall contact intensity by reported by adult respondents is the same as the mean number of overall close contacts reported by adult respondents (graph a). Contact numbers between child ‘respondents’ and contacts in each age group were estimated from data on contact between adult respondents and child contacts. Ranges shown are bootstrapped 95% plausible ranges.

**Figure 4.**
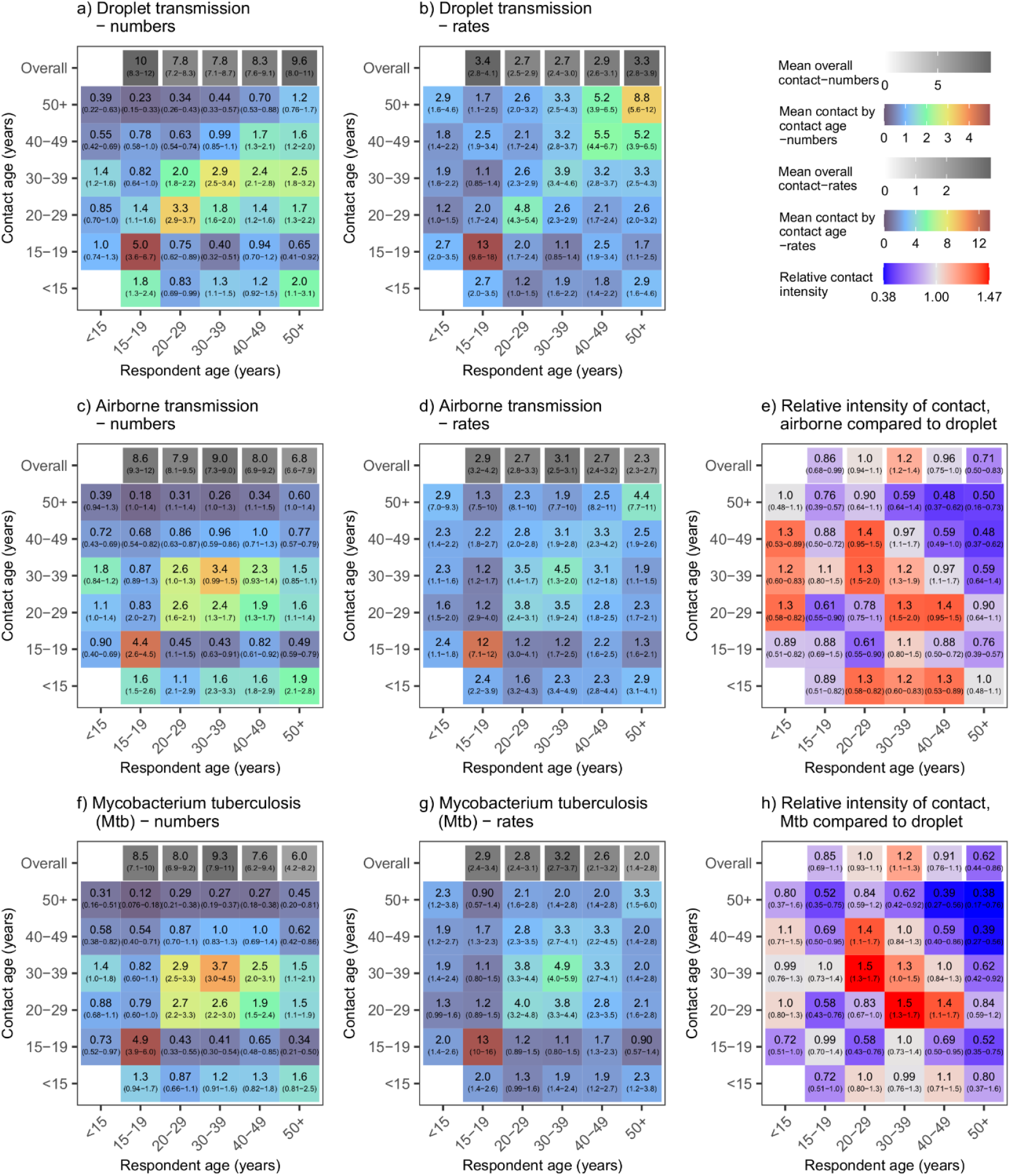
Age mixing matrices relevant for droplet transmission, non-saturating airborne transmission, and *Mycobacterium tuberculosis* transmission, for Western Cape. Graphs a, c, and f show absolute contact intensities between respondents and contacts in each age group. Graphs b, d, and g show intensities of contact between each member of each age group. Graphs e and h show intensities for airborne infections and *Mtb* compared to intensities for droplet infections respectively. Numbers shown in graph a are the mean number of contacts respondents in each age group have with contacts in each age group per day. Numbers shown in graph b are the rate of contact between each individual in the population per day, expressed as rates ×10^5^. ‘Numbers’ and ‘rates’ in graphs c, d, f, and g are standardised so that the mean overall contact intensity by reported by adult respondents is the same as the mean number of overall close contacts reported by adult respondents (graph a). Contact numbers between child ‘respondents’ and contacts in each age group were estimated from data on contact between adult respondents and child contacts. Ranges shown are bootstrapped 95% plausible ranges.

Estimated contact patterns by age did not vary greatly between the three transmission routes/infections, in either setting. Age mixing patterns were less assortative, however, in the non-saturating airborne and *Mtb* matrices compared to the droplet matrices, in both settings. The exception to this was contact between 15-19 year olds in KwaZulu-Natal, which was more intense in the non-saturating airborne and *Mtb* matrices than the droplet matrices. In both communities, relative to other adult age groups, overall contact intensities were lower in 50+ year olds when considering contact relevant for non-saturating airborne transmission or the transmission of *Mycobacterium tuberculosis* than when considering contact relevant for droplet transmission.

## Discussion

The majority of data used to generate age-mixing matrices used in transmission models is close contact data – data on contacts involving a face-to-face conversation. For infections where airborne transmission is common, close contact data is likely to miss many potential effective contacts. Using data from two provinces in South Africa, we created improved estimates of age-mixing patterns for airborne infections, using a wider ‘casual contact’ definition that incorporated anybody ‘sharing space’ indoors. We also demonstrated a novel method for generating age-mixing matrices relevant for the transmission of Mtb, an airborne infection where long disease durations and the saturation of household contacts play an important role in transmission dynamics. Finally, we estimated age-mixing patterns from close contact data using the most commonly used method, which we have labelled as relevant for droplet transmission. In our settings, contact patterns did not vary greatly between contacts relevant for droplet infections and those relevant for non-saturating airborne transmission or Mtb. Using close contact data in models of the transmission of Mtb or other vairborne infections in our study communities, however, may mean that the importance of adults aged 50+ to transmission is overestimated.

Very few data are available on casual contact patterns. Previous studies in the same community in Western Cape have found greater drops in casual contact time than in close contact numbers in older age groups(6), and decreases in indoor casual contact numbers with age(7). Another study in the same community found high levels of age-assortative mixing with respect to casual contact time in schools and workplaces(8). More data are needed on casual contact patterns, and age-mixing patterns in particular, in order to determine whether the findings of this study are generalisable to other settings, and to improve the predictions from mathematical models of the transmission of *Mtb* and other airborne infections.

Our approaches to generating the separate droplet and airborne transmission matrices are necessarily simplifications, and many infections will not fit neatly into these two categories. There is considerable uncertainty about the role of different transmission routes to the spread of many infections, and both airborne and droplet transmission are thought to play a role in the transmission of some respiratory infections, including SARS-CoV-2(9). For these infections, an intermediate matrix may be preferable.

There are two main differences between our droplet and airborne/*Mtb* age-mixing matrices. The first is the type of non-household contacts considered: close (face-to-face conversation) or casual (sharing space indoors) respectively. The second is that the airborne and (non-household component of the) *Mtb* matrices are based on contact time, rather than unique contact numbers. The primary reason for using contact time for casual contacts is that respondents are unlikely to be able to estimate unique casual contact numbers for many locations they visit, necessitating the use of contact time, or assumptions about the rate of turnover of unique people in a location. For our droplet transmission matrices, we chose to use unique contact numbers in a 24-hour period, as that is the most commonly used method, and therefore allows comparisons to be made with what is typically done. It should be noted, however, that both the choice of a 24-hour time period, and the lack of any weighting or restrictions by contact duration or other measures of closeness, are relatively arbitrary choices.

Robust evidence as to the types of contact most relevant to transmission are limited for respiratory infections. A number of studies have compared the fit to data on varicella, parvovirus B19, or influenza A seroprevalence by age of models parametrised using contact patterns generated from close contact data in a range of different ways(10-12). Overall, these studies suggest that analysis methods that give greater weight to more intimate contacts may be preferable in some circumstances. This could be achieved by restricting what counts as a contact to those involving physical touch and/or a minimum contact duration, or by using contact time rather than contact numbers. It has been suggested that approaches based on contact numbers may be more suitable for more highly transmissible infections, where only a short duration of contact is needed for transmission, whereas approaches based on contact time may be more suitable for less transmissible infections, where repeated or longer contacts are needed(13).

Fewer studies have considered expanding the pool of contacts beyond close contacts only, to also include casual contacts. One study however, that had paired individual-level contact data and pandemic influenza A serological data, found that models that included a variable for number of locations visited were strongly supported over those that only included variables for age and close contact numbers(14). This suggests that airborne transmission may play a role in the spread of influenza A, and/or that the standard close contact definition misses a significant proportion of contacts at risk of droplet transmission.

Other factors may also influence airborne and *Mtb* transmission risk, which are not accounted for in the analyses. Ventilation rates play a large role in determining airborne infection risk(15), and weighting by ventilation rates would improve our airborne and *Mtb* matrices. Unfortunately, few data on ventilation rates by location type are available, and they show large amounts of variation between locations, and between the same location on different days(16). Saturation of contacts may occur for infections other than *Mtb*, particularly highly transmissible pathogens such as measles virus. An approach based on casual contact numbers may be preferable for these infections, but would be highly dependent on assumptions made about how unique contact numbers are related to estimates of cross-sectional numbers of people present.

There are a number of limitations when using casual contact data to estimate mixing patterns. Firstly, estimates of contact time in places where large numbers of people are present are likely to be less reliable. This is because people’s estimates of the number of people present are likely to be poor, and because the assumption that there is a risk of transmission between all people present in the space may not be true in larger spaces. In our main analysis, when estimating contact time, we cap the number of people at risk of transmission at 100. In our sensitivity analyses, we show that using a cap of 20 people, or not capping the numbers of people, has a moderate impact on casual contact time age mixing matrices (see Appendix). Conducting similar sensitivity analyses may be necessary when using age mixing matrices calculated using casual contact time in mathematical models

A second limitation is that the approach we use to determining the ages of adults present in locations other than respondents’ own homes is indirect, and relies on the assumption that the age distribution of adults present in a location type reflects the duration of time respondents of different ages reported spending in that location type. This may not always be a reasonable assumption, if different age groups tend to visit different locations of the same type (or at different times), or substantial mixing occurs with people from outside the study setting. These issues are discussed further in McCreesh *et al* 2019*(6)*.

An additional limitation of our estimates for KwaZulu-Natal only is that we did not recruit 15-17 years olds, and instead assumed in the analysis that contact by 18-19 year olds was representative of contact by all 15-19 year olds. This is unlikely to be true, with contacts by 15-17 and 18-19 year olds differing greatly in Western Cape (see Appendix, Figure S7). For this reason, our estimates for 15-19 year olds should be treated with caution for KwaZulu-Natal.

To conclude, overall our estimated age-mixing matrices for droplet transmission, non-saturating airborne transmission, and *Mtb* were not substantially different from each other for either setting. This provides some reassurance that the widespread use of close contact data to parameterise age-mixing matrices for transmission models of airborne infections may not be resulting in major inaccuracies. Some differences were observed however, particularly in the oldest age group, and our data were from two South African settings only. We recommend that future social contact surveys should collect data on casual contacts as well as close contacts, to determine whether the similarity between different types of contact pattern is true across other settings. We would also urge mathematical modellers to consider whether unique close contact numbers in a 24-hour period are the most appropriate contacts for the infection and scenario they are simulating, and to consider performing sensitivity analyses when there is uncertainty as to the most appropriate contact definition.

## Data Availability

Analysis code and data are available from https://github.com/NickyMcC/CasualAgeMixing

## Acknowledgements

This research is jointly funded by the UK Medical Research Council (MRC) and the UK Department for International Development (DFID) under the MRC/DFID Concordat agreement MR/P002404/1. The support of the Economic and Social Research Council (IK) is gratefully acknowledged. The project is partly funded by the Antimicrobial Resistance Cross Council Initiative supported by the seven research councils in partnership with other funders including support from the GCRF. Grant reference: ES/P008011/1. NM is additionally funded the Wellcome Trust (218261/Z/19/Z). RGW is funded by the Wellcome Trust (218261/Z/19/Z), NIH (1R01AI147321-01), EDTCP (RIA208D-2505B), UK MRC (CCF17-7779 via SET Bloomsbury), ESRC (ES/P008011/1), BMGF (OPP1084276, OPP1135288 & INV-001754), and the WHO (2020/985800-0).

We would like to thank the social contact survey respondents, and all of the fieldworkers involved in the data collection (In KwaZulu-Natal: Nkosingiphile Buthelezi, Zilethile Khumalo, Sifundesihle Malembe, Zodwa Mkwanazi, Sanele Mthiyane. In Western Cape: Sinayo Dyubhele, Mzukisa Diniso, Olwethu Kemele, Zilungile Majola Nyembezi, Nomonde Vungama & Nontyatyambo Bangani).

## Data/code availability

Analysis code and data are available from https://github.com/NickyMcC/CasualAgeMixing

## References

1. Prem K, Cook AR, Jit M. Projecting social contact matrices in 152 countries using contact surveys and demographic data. PLOS Computational Biology. 2017;13(9):e1005697.

2. Van Hoang T, Coletti P, Melegaro A, Wallinga J, Grijalva C, Edmunds J, et al. A systematic review of social contact surveys to inform transmission models of close contact infections. bioRxiv. 2018:292235.

3. Raffalli J, Sepkowitz KA, Armstrong D. Community-based outbreaks of tuberculosis. Archives of Internal Medicine. 1996 May 27;156(10):1053–60.

4. Ku C-C, MacPherson P, Khundi M, Nzawa R, Feasey HR, Nliwasa M, et al. Estimated durations of asymptomatic, symptomatic, and care-seeking phases of tuberculosis disease. medRxiv. 2021:2021.03.17.21253823.

5. McCreesh N, White RG. An explanation for the low proportion of tuberculosis that results from transmission between household and known social contacts. Scientific reports. 2018;8(1):5382.

6. McCreesh N, Morrow C, Middelkoop K, Wood R, White RG. Estimating age-mixing patterns relevant for the transmission of airborne infections. Epidemics. 2019;28:100339.

7. Wood R, Racow K, Bekker L-G, Morrow C, Middelkoop K, Mark D, et al. Indoor social networks in a South African township: potential contribution of location to tuberculosis transmission. PLoS One. 2012;7(6):e39246.

8. Andrews JR, Morrow C, Walensky RP, Wood R. Integrating social contact and environmental data in evaluating tuberculosis transmission in a South African township. Journal of Infectious Diseases. 2014;210(4):597–603.

9. Greenhalgh T, Jimenez JL, Prather KA, Tufekci Z, Fisman D, Schooley R. Ten scientific reasons in support of airborne transmission of SARS-CoV-2. The lancet. 2021;397(10285):1603–5.

10. Ogunjimi B, Hens N, Goeyvaerts N, Aerts M, Van Damme P, Beutels P. Using empirical social contact data to model person to person infectious disease transmission: an illustration for varicella. Mathematical biosciences. 2009;218(2):80–7.

11. Melegaro A, Jit M, Gay N, Zagheni E, Edmunds WJ. What types of contacts are important for the spread of infections? Using contact survey data to explore European mixing patterns. Epidemics. 2011;3(3-4):143-51.

12. Kucharski AJ, Kwok KO, Wei VWI, Cowling BJ, Read JM, Lessler J, et al. The Contribution of Social Behaviour to the Transmission of Influenza A in a Human Population. PLOS Pathogens. 2014;10(6):e1004206.

13. Iozzi F, Trusiano F, Chinazzi M, Billari FC, Zagheni E, Merler S, et al. Little Italy: An Agent-Based Approach to the Estimation of Contact Patterns-Fitting Predicted Matrices to Serological Data. PLOS Computational Biology. 2010;6(12):e1001021.

14. Kwok KO, Cowling BJ, Wei VW, Wu KM, Read JM, Lessler J, et al. Social contacts and the locations in which they occur as risk factors for influenza infection. Proceedings of the Royal Society B: Biological Sciences. 2014;281(1789):20140709.

15. Li Y, Leung GM, Tang J, Yang X, Chao C, Lin JZ, et al. Role of ventilation in airborne transmission of infectious agents in the built environment-a multidisciplinary systematic review. Indoor air. 2007;17(1):2–18.

16. Taylor JG, Yates TA, Mthethwa M, Tanser F, Abubakar I, Altamirano H. Measuring ventilation and modelling M. tuberculosis transmission in indoor congregate settings, rural KwaZulu-Natal. Int J Tuberc Lung Dis. 2016 Sep;20(9):1155–61.

