## Appendix for "Improving estimates of social contact patterns for the airborne transmission of respiratory pathogens"

**Appendix: Improving estimates of social contact patterns for the airborne transmission of *Mycobacterium tuberculosis* and other respiratory infections**

### Methods

#### Weighting

All analyses of contact numbers and contact time were weighted. For KwaZulu-Natal, they were weighted to the study population composition by age group (18-19, 20-29, 30-39, 40-49, 50+) and sex. For Western Cape, they were weighted to the study population composition by age group (15-17, 18-19, 20-29, 30-39, 40-49, 50+) and sex. As fewer respondents were asked about Fridays and Saturdays, the Western Cape data were also weighted by the day of the week.

#### Estimating close contact numbers

Overall close contact numbers by respondent characteristic were estimated using the total number of close contacts that the respondent reported, and the number of those contacts who were household members. Respondents who reported more household contacts than overall contacts were excluded from the analysis.

Close contact age mixing matrices were generated using data on close contacts on which more detailed information were available (all contacts if respondents reported ≤10, a random 10 if they reported more). When generating the central estimate, contact numbers by age group for respondents were multiplied by $T / D_a$, where T was the total number of contacts that they reported, and $D_a$ was the total number of contacts whose age was provided. Respondents who reported a non-zero number of close contacts, but who did not give the age of any of their contacts were excluded. 95% plausible intervals were generated using 10,000 bootstrapped samples, re-sampling respondents with replacement within age categories, and re-sampling *T* contacts with replacement from the set of all contacts of the respondent on which detailed information were collected(1). Only sampled contacts who were or weren't reported to be a member of the respondent's household were included when estimating household and 'other' age-mixing patterns respectively.

The age mixing matrices were adjusted to be symmetrical, using the study community age structures. Data on adult contacts with children were used to estimate child contact with adults. To allow comparison between the two study communities, and between close and casual age mixing patterns, the lowest respondent age group was set at 15-19 years for both surveys. As 15-17 year olds were not interviewed in KwaZulu-Natal, we assumed that contact patterns in 18-19 years olds were representative of contact patterns in all 15-19 year olds, and adjusted the weights accordingly.

#### Estimating close contact time

The approach used for estimating close contact time was the same as that used for estimating close contact numbers, except that contact numbers were multiplied by the amount of time that respondents reported spending with each contact that day.

Contacts with the contact duration missing were excluded when generating bootstrap samples for estimating close contact time age mixing patterns. When generating the central estimate, contact numbers by age group for respondents were multiplied by $T / D_{ad}$, where T was the total number of contacts that they reported, and $D_{ad}$ was the total number of contacts whose age and duration were both provided.

#### Estimating casual contact time

For each location visited, respondents were asked to identify the location type, from a list of frequently visited location types identified by local researchers and fieldworkers before the start of data collection. If the interviewer could not identify the correct location type on the list, they selected 'Other' and gave details. In the analysis, locations were excluded if it was considered likely that most all or of the time would have been spent outdoors (e.g. if the details given were 'gardening'). A number of responses were re-coded, if it was considered plausible from the details given that the location was covered by one of the original options (e.g. 'domestic worker' was changed to 'House off plot'). A number of new location categories were added, if reported by multiple respondents (e.g. 'factory'). Finally, remaining responses in the 'Other' category were recorded as 'Other' if the type of location could be determined from the free text variable, and 'Missing' if it could not be.

Respondents were excluded from all casual contact time analyses if:

1) They reported visiting no locations (including their own home) and using no transport
2) The variable giving the total number locations visited was missing, and no information was provided on any locations visited
3) No information was available on any of the locations visited or transport used

Casual contact time was estimated as the duration of time that respondents reported spending in a location, multiplied by the number of people that they estimated were present at the location, halfway through the time that they were there. In the analysis, total numbers of people present were capped at a maximum of 100, as above this value, it is unlikely that the respondent had sufficient contact with each person present to allow transmission. Estimates of numbers of adults and children present were reduced by the same proportion for each location, to give a maximum total number of people present of 100.

If the estimated number of people or children present was missing for a location, or if the estimated number of children present was greater than the estimated total number of people present, then the numbers of adults and children present were set equal to the mean reported number of adults and children present at locations of that type in the same community (KwaZulu-Natal or Western Cape). These numbers were rounded to the nearest whole number when generating bootstrap samples. If the duration of time spent in the location was missing, the duration was set equal to the mean duration for locations of that type.

Age mixing matrices for casual contact time were generated using the data on locations visited, the duration of time spent in the location, and the estimated number of adults and children present. Central estimates were generated using the method outlined in McCreesh *et al*(2). As data were collected on numbers of people and children present in indoor locations only, and not on the ages of adults present, the age distribution of adult casual contacts needed to be estimated. To do so, we assumed that the age distribution of adults present in each location type matched the age distribution of respondents who reported visiting locations of that type, weighted by the duration of time they reported spending in that location type and weighted to the sampled population age and sex distribution. To generate plausible ranges, 10,000 bootstrap samples were generated, re-sampling respondents with replacement within age categories, and sampling the ages of adults present by resampling with replacement respondents who reported visiting locations of that type (weighted by duration, and to the sampled age and sex distribution). The number of children present was set equal to number of children present reported by the respondent. Contact times were then estimated by multiplying the duration of time the respondent reported spending in each location by the sampled number of people of each age group present.

The age mixing matrices were made symmetrical, using data on adult contact time with children to estimate child contact time with adults. As respondents were asked to estimate numbers of children present who were aged under 15 years, we set the lowest respondent age group to be 15-19 years. As 15-17 year olds were not interviewed in KwaZulu-Natal, we assumed that contact patterns in 18-19 years olds were representative of contact patterns in all 15-19 year olds, and adjusted the weights accordingly.

#### Estimating age-mixing patterns relevant for the transmission of droplet infections

Age-mixing patterns relevant for the transmission of droplet infections were assumed to be equal to age mixing patterns calculated from close contact numbers.

#### Estimating age-mixing patterns relevant for the transmission of airborne infections

To generate age-mixing patterns relevant for the transmission of airborne infections, we summed estimated close contact time between household members, and estimated casual contact time occurring in locations other than the respondents' own houses. 95% plausible ranges were generated by pairing each of the 10,000 close contact household bootstrapped matrices with one of the 10,000 outside-household casual contact time bootstrapped matrices.

To allow direct comparisons to be made between the different age mixing matrices, the matrices for airborne infections (central estimate and 10,000 individual bootstrapped matrices) were then adjusted to give the same mean contact intensity between adults as the matrices for close contacts (using the central estimate matrices and 10,000 individual bootstrapped matrices respectively).

#### Estimating age-mixing patterns relevant for the transmission of *Mycobacterium tuberculosis*

It is estimated that, in high incidence settings, only 8-19% of tuberculosis comes from household transmission(2). Long durations of disease(3) also mean that transmission to household members may partially saturate, meaning that the relationship between contact time and transmission is non-linear for household contacts. We therefore estimated age mixing matrices relevant to the transmission of *Mtb* by creating weighted averages of close contact numbers with household members, and casual contact time occurring outside respondents' own households.

For each pair of bootstrapped household close contact number and non-household casual contact time matrices, a proportion of contact that should occur in households was sampled from a uniform distribution between 8-16% (with 12% used for the central estimate). A weighted average was then generated, given the desired proportion of overall 'contact' by adults occurring in households.

To allow direct comparisons to be made between the different age mixing matrices, the matrices for *Mtb* (central estimate and 10,000 individual bootstrapped matrices) were then adjusted to give the same mean contact intensity between adults as the matrices for close contacts (using the central estimate matrices and 10,000 individual bootstrapped matrices respectively).

##### Sensitivity analyses

In our main analysis, we cap the number of people at risk of infection in locations (i.e. casual contact numbers) at a maximum of 100. In the sensitivity analysis, we explore the effect of setting the cap at 20, or not having a cap.

When generating the non-saturating airborne and *Mtb* age-mixing matrices, we used close contact time and close contact number data respectively to estimate contact time between household members. In the sensitivity analysis, we explore the effect of using casual contact time.

For all sensitivity analysis age-mixing matrices, we rescaled contact times to give the same overall mean contact hours per adult in the sensitivity analysis as in the main analysis. This was done because the primary use of age mixing matrices is in mathematical modelling, where it is usually the relative values of the cells in the matrices that has an impact on model dynamics, not the absolute values.

#### Results

##### Missing/incomplete data

###### Close contact data

- The reported number of household contacts was higher than the reported total number for two respondents in KwaZulu-Natal. They were excluded from all analyses of close contacts.
- Three respondents in KwaZulu-Natal had the contact age missing for all of their contacts, and were excluded from the age-mixing analysis.
- The number of household contacts was unknown for 11 respondents in KwaZulu-Natal. They were not included in the analysis of household or non-household contacts (Figures S8-S11).
- Contact ages were missing for 54 contacts in KwaZulu-Natal and 16 contacts in Western Cape

- Whether a contact is a household member was missing for 16 contacts in KwaZulu-Natal and two in Western Cape
- Contact durations were missing for 16 contacts in KwaZulu-Natal and three contacts in Western Cape. No respondents were missing the duration of time for all contacts.

##### Casual contact data

- One person in KwaZulu-Natal and eight in Western Cape reported that they visited no locations (including their own house) and used no transport used. This was considered to be implausible, and they were excluded from the casual contact time analyses.
- The 'total number of locations visited' variable was missing for four respondents in Western Cape, and they gave no information on any locations visited. They were excluded from the casual contact time analyses.
- 43 respondents in Western Cape reported the total number of locations visited, but did not give any information on any of the locations visited. They did not differ from other respondents in Western Cape in respect to the reported number of locations visited (no details given: mean locations = 3.0, 95% CI 2.5-3.5, detail given: 3.0 (2.9-3.1), and were excluded from the casual contact time analyses.
- The number of people and/or number of children present was missing for 102 locations in KwaZulu-Natal and 220 in Western Cape. The reported number of children present was greater than reported total number of people present for seven locations in KwaZulu-Natal and none in Western Cape.
- The duration of time spent in a location was missing for two locations in Western Cape and none in KwaZulu-Natal.

#### Sensitivity analyses

##### Changing the cap on people at risk in locations

Overall casual contact time was lower when the number of people at risk was capped at 20, and higher when it was not capped, compared to when it was capped at 100 people (Figure S1). Changing the cap had a moderate effect on casual contact time age-mixing patterns, although most changes were not large compared to the breadths of the 95% plausible ranges (Figure S2).

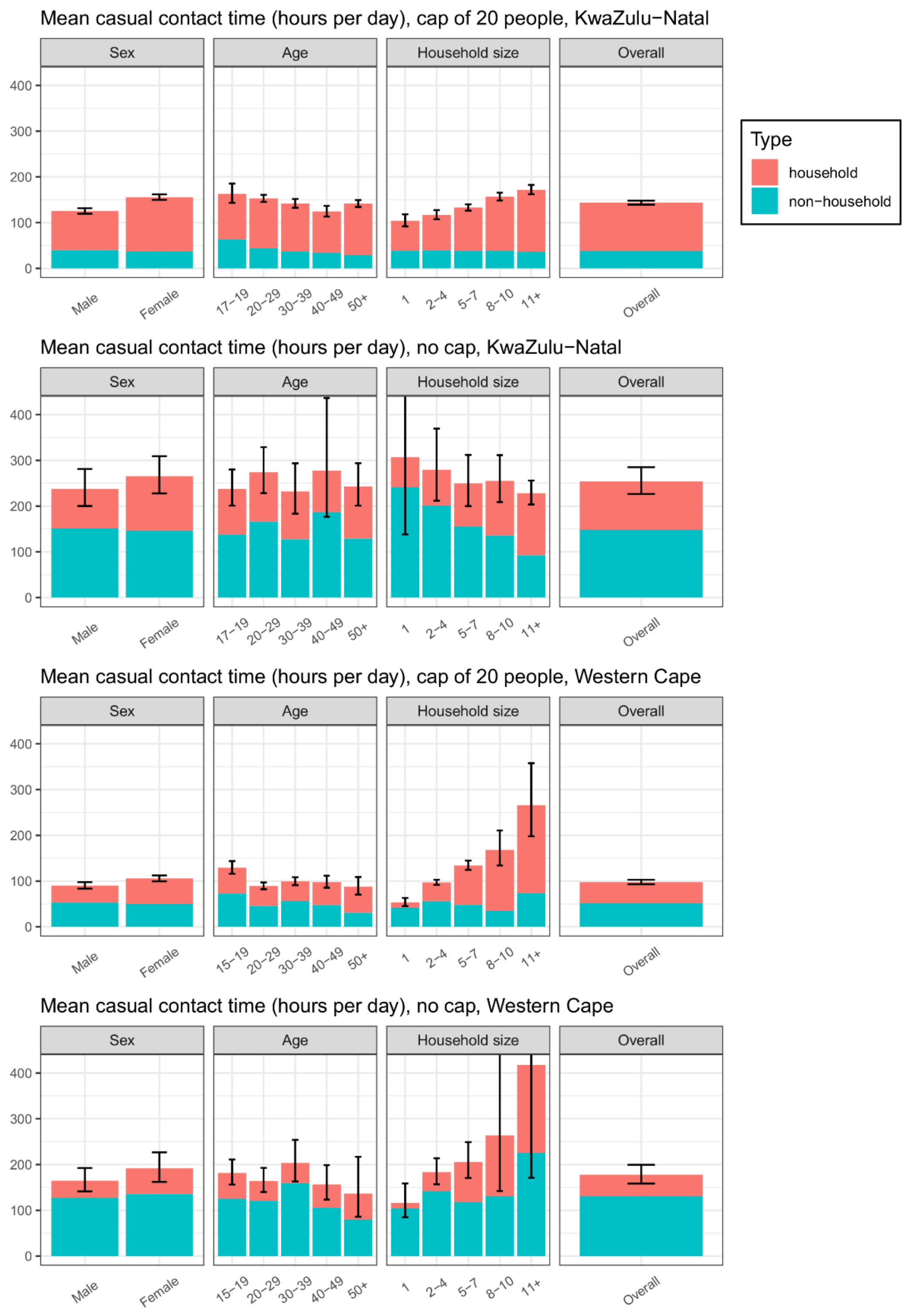


Figure S1. Household and non-household close contact numbers, close contact time, and casual contact time in KwaZulu-Natal and Western Cape, with a cap of 20 people at risk per location, and with no cap on the number at risk. Error bars show 95% confidence intervals for total contact numbers or time.

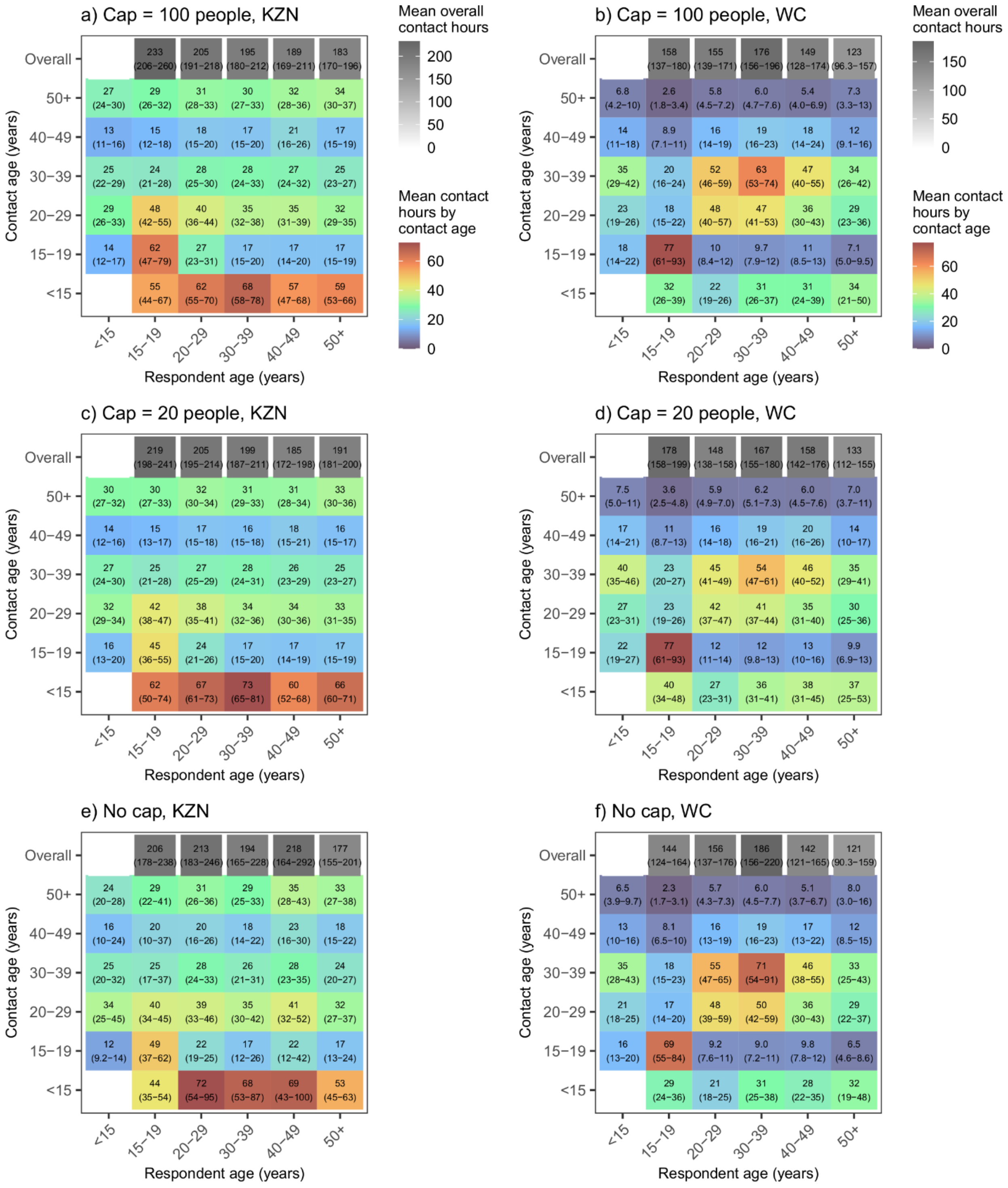


Figure S2. Casual contact time age mixing patterns for KwaZulu-Natal and Western Cape with a cap of 100 people at risk per location (baseline scenario), a cap of 20 people (sensitivity analysis), and no cap (sensitivity analysis). Figures a and b show estimated mean contact hours per day between respondents and contacts in each age group. Figures c-f show estimated mean contact hours, scaled to give the same overall mean adult contact hours per adult as figures a (c and e) and b (d and f).

#### Generating age-mixing matrices using casual contact time for household contact

Calculating age-mixing matrices relevant for non-saturating airborne and *Mtb* transmission using casual contact time data for contact in between household members had very little effect on estimated age-mixing patterns (Supporting information, Figures S3 and S4). The exception to this was contact relevant to airborne transmission between 15-19 year olds in KwaZulu-Natal, which was lower when casual contact time data were used than in the main analysis.

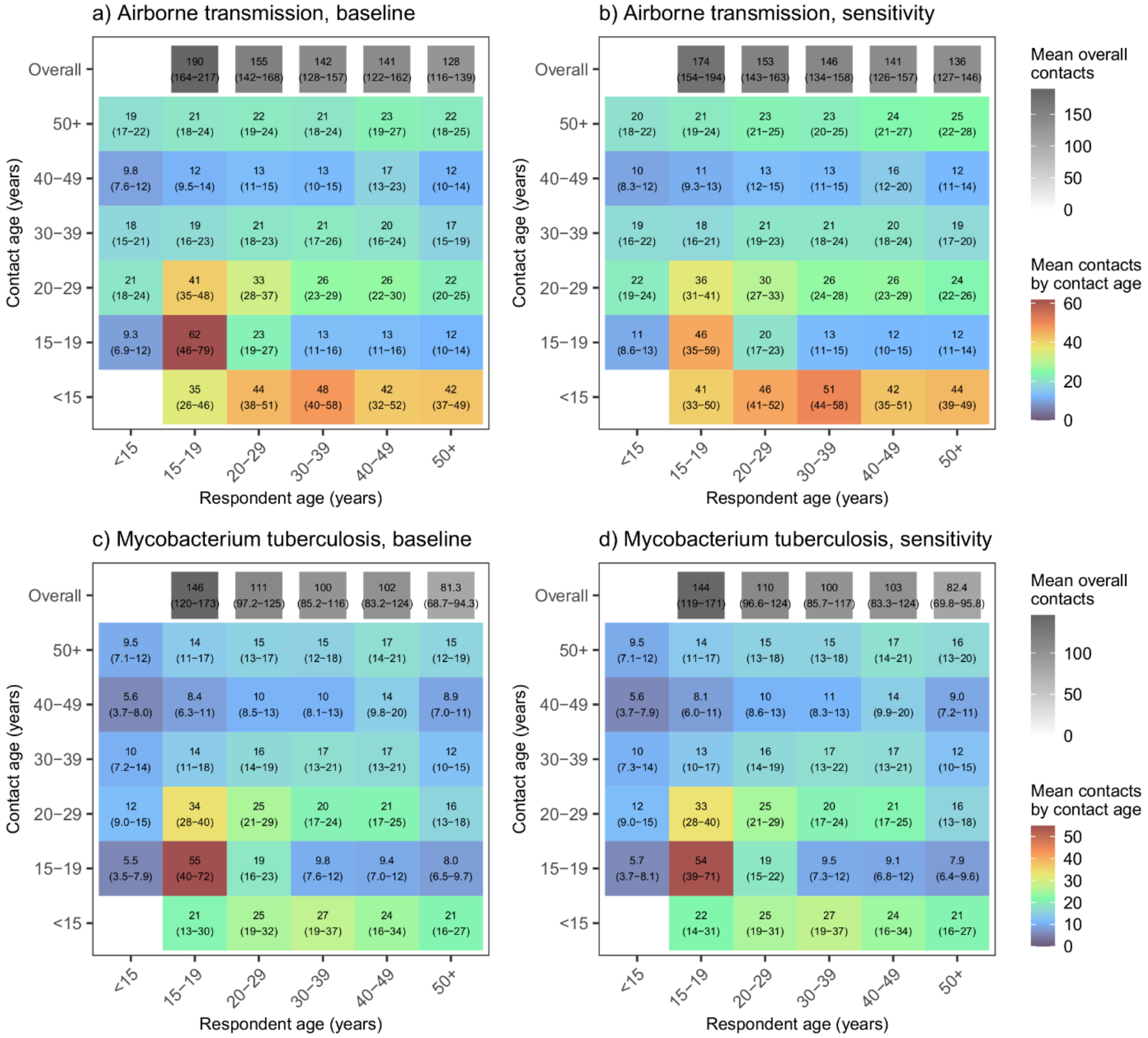

Figure S3. Age mixing patterns relevant for non-saturating airborne transmission and the transmission of *Mycobacterium* tuberculosis in KwaZulu-Natal, with household age mixing estimated using close (baseline) and casual (sensitivity analysis) contact data. Figures show mean contact hours per day between respondents and contacts in each age group. Numbers in b and d are scaled to give the same overall mean adult contact hours per adult as figures a and c respectively.

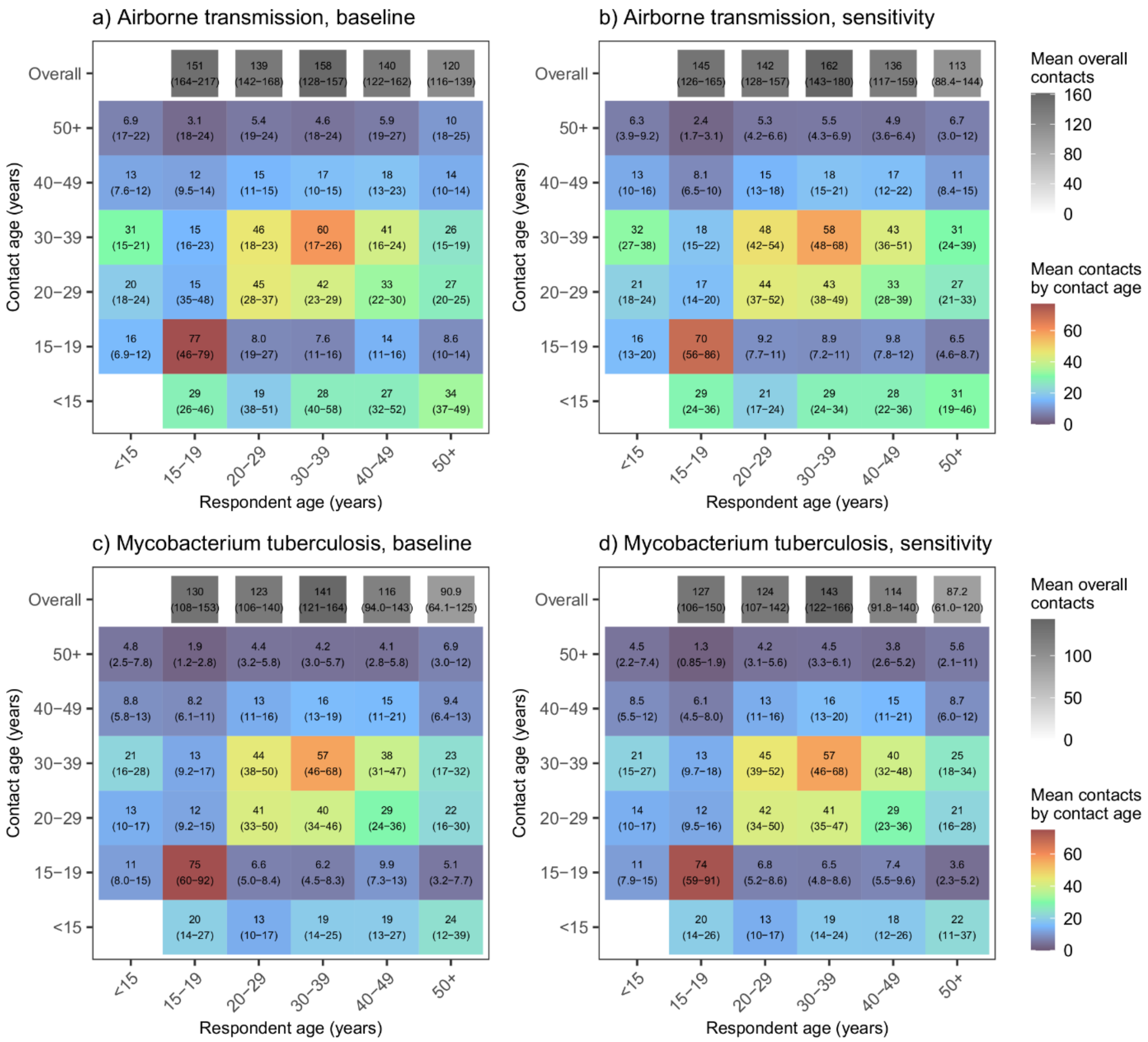


Figure S4. Age mixing patterns relevant for non-saturating airborne transmission and the transmission of *Mycobacterium* tuberculosis in Western Cape, with household age mixing estimated using close (baseline) and casual (sensitivity analysis) contact data.  Figures show mean contact hours per day between respondents and contacts in each age group. Numbers in b and d are scaled to give the same overall mean adult contact hours per adult as figures a and c respectively.

#### Additional results tables and figures

##### Close contact numbers, close contact time, and casual contact time tables

**Close contact numbers - KwaZulu-Natal**

| | | Overall | | Household members | | Non-household members | |
|---|---|---|---|---|---|---|---|
| | | Mean (95% CI) | p-value | Mean (95% CI) | p-value | Mean (95% CI) | p-value |
| **Sex** | Male | 7.0 (6.6-7.4) | <0.001 | 3.8 (3.6-4.0) | <0.001 | 3.2 (2.8-3.5) | <0.001 |
| | Female | 7.6 (7.3-8.0) | | 5.1 (4.9-5.3) | | 2.5 (2.2-2.9) | |
| **Age** | 15-19 | 7.9 (7.0-9) | 0.4 | 4.9 (4.3-5.5) | 0.3 | 2.9 (2.1-3.8) | 1.0 |
| | 20-29 | 8.2 (7.6-8.7) | | 4.8 (4.5-5.1) | | 3.3 (2.9-3.9) | |
| | 30-39 | 7.4 (6.9-8.0) | | 4.5 (4.2-4.9) | | 2.9 (2.5-3.4) | |
| | 40-49 | 6.5 (6.0-7.0) | | 4.2 (3.8-4.6) | | 2.4 (2.0-2.8) | |
| | 50+ | 6.8 (6.3-7.4) | | 4.5 (4.2-4.8) | | 2.3 (1.8-2.8) | |
| **Residence** | Peri-Urban | 7.0 (6.6-7.5) | 0.6 | 4.3 (4.0-4.5) | 0.001 | 2.7 (2.4-3.1) | 0.08 |
| | Urban | 7.6 (7.3-8.0) | | 5.0 (4.8-5.2) | | 2.6 (2.4-2.9) | |

| | | | | | | | |
|---|---|---|---|---|---|---|---|
| | Rural | 7.4 (6.0-9.1) | | 3.5 (3.1-3.9) | | 3.9 (2.7-5.8) | |
| **Monthly household income** | Less than R1000 | 6.8 (6.3-7.4) | 0.4 | 4.2 (4.0-4.5) | 0.7 | 2.5 (2.1-3.1) | 0.2 |
| | R1000 - R2500 | 7.6 (7.2-7.9) | | 5.0 (4.8-5.3) | | 2.6 (2.3-2.9) | |
| | R2500 - R5000 | 7.2 (6.5-7.9) | | 4.2 (3.8-4.5) | | 3.1 (2.5-3.8) | |
| | R5000 - R10000 | 7.1 (6.3-8.0) | | 4.2 (3.7-4.9) | | 2.9 (2.3-3.6) | |
| | More than R10000 | 9.7 (7.5-13) | | 4.6 (3.8-5.7) | | 5.1 (3-8.6) | |
| | Unknown/missing | 5.5 (4.3-7.0) | | 2.5 (1.8-3.4) | | 3.0 (2.0-4.3) | |
| **Employment** | Full-time | 7.4 (6.7-8.2) | 0.9 | 3.8 (3.5-4.2) | <0.001 | 3.6 (2.9-4.4) | 0.002 |
| | Part-time | 7.1 (6.1-8.2) | | 4.0 (3.3-4.8) | | 3.0 (2.4-3.8) | |
| | None/missing | 7.4 (7.1-7.6) | | 4.8 (4.6-5.0) | | 2.5 (2.3-2.8) | |
| **Household size** | 1 | 5.5 (4.9-6.2) | <0.001 | 2.9 (2.5-3.3) | <0.001 | 2.6 (2.1-3.3) | 0.3 |
| | 2-4 | 6.2 (5.5-7.0) | | 3.3 (2.9-3.6) | | 3.0 (2.3-3.8) | |
| | 5-7 | 7.2 (6.7-7.7) | | 4.2 (4.0-4.4) | | 3.0 (2.6-3.6) | |
| | 8-10 | 7.7 (7.1-8.3) | | 5.2 (4.9-5.4) | | 2.5 (2.1-3.0) | |
| | 11+ | 8.5 (8.0-9.0) | | 5.8 (5.4-6.2) | | 2.6 (2.3-3.0) | |
| **Day reported** | Sunday | 7.6 (6.8-8.4) | 0.1 | 4.5 (4.1-4.9) | 0.3 | 3.2 (2.5-4.0) | 0.001 |

| | | | | |
|---|---|---|---|---|
| | Monday | 7.5 (6.7-8.4) | 4.6 (4.2-5.1) | 2.9 (2.2-3.8) |
| | Tuesday | 6.9 (6.4-7.4) | 4.8 (4.4-5.2) | 2.0 (1.7-2.4) |
| | Wednesday | 7.2 (6.7-7.7) | 4.6 (4.2-5.0) | 2.6 (2.2-3.0) |
| | Thursday | 8.1 (7.3-9.1) | 4.6 (4.2-5.1) | 3.4 (2.7-4.4) |
| | Friday | 7.0 (6.5-7.7) | 4.5 (4.1-4.9) | 2.6 (2.1-3.1) |
| | Saturday | 7.2 (6.5-7.9) | 4.5 (4.1-4.9) | 2.7 (2.2-3.4) |
| **Total** | | 7.4 (7.1-7.6) | 4.6 (4.4-4.7) | 2.8 (2.5-3.0) |

Table S1. Mean overall, household, and non-household close contact numbers in KwaZulu-Natal.

**Close contact numbers – Western Cape**

| | | Overall | | Household members | | Non-household members | |
|---|---|---|---|---|---|---|---|
| | | Mean (95% CI) | p-value | Mean (95% CI) | p-value | Mean (95% CI) | p-value |
| **Sex** | Male | 7.9 (7.3-8.4) | <0.001 | 1.8 (1.7-2.0) | <0.001 | 6.0 (5.5-6.6) | <0.001 |

| | | | | | | | |
|---|---|---|---|---|---|---|---|
| | Female | 8.3 (7.6-9.2) | | 2.6 (2.5-2.8) | | 5.7 (5.0-6.6) | |
| **Age** | 15-19 | 11 (9.3-13) | <0.001 | 3.1 (2.8-3.5) | <0.001 | 8.0 (6.3-10) | 0.02 |
| | 20-29 | 7.8 (7.2-8.4) | | 1.9 (1.7-2.1) | | 5.9 (5.3-6.5) | |
| | 30-39 | 7.7 (6.8-8.7) | | 2.0 (1.8-2.2) | | 5.7 (4.8-6.7) | |
| | 40-49 | 7.8 (7.0-8.6) | | 2.6 (2.3-3.0) | | 5.2 (4.4-6.0) | |
| | 50+ | 8.7 (6.7-11) | | 2.9 (2.4-3.5) | | 5.8 (3.8-8.7) | |
| **Residence** | Peri-Urban | 8.1 (7.6-8.6) | | 2.2 (2.1-2.3) | | 5.9 (5.4-6.4) | |
| | Urban | NA | | NA | | NA | |
| | Rural | NA | | NA | | NA | |
| **Monthly household income** | Less than R1000 | 9.2 (6.5-13) | 0.7 | 2.0 (1.7-2.5) | 0.4 | 7.2 (4.6-11) | 0.6 |
| | R1000 - R2500 | 7.8 (7.1-8.6) | | 2.1 (1.8-2.3) | | 5.7 (5.1-6.5) | |
| | R2500 - R5000 | 8.6 (8.0-9.3) | | 2.2 (2-2.4) | | 6.4 (5.8-7.1) | |
| | R5000 - R10000 | 7.2 (6.3-8.2) | | 2.0 (1.8-2.3) | | 5.2 (4.3-6.2) | |
| | More than R10000 | 8.5 (6.7-11) | | 2.7 (2.3-3.2) | | 5.8 (4.1-8.2) | |
| | Unknown/missing | 7.1 (6.2-8.1) | | 2.7 (2.4-3.1) | | 4.4 (3.5-5.4) | |
| **Employment** | Full-time | 8.3 (7.7-9.0) | 0.1 | 2.0 (1.9-2.2) | <0.001 | 6.3 (5.7-7.0) | 0.009 |

| | | | | | | | |
|---|---|---|---|---|---|---|---|
| | Part-time | 8.5 (7.1-10) | | 1.9 (1.7-2.2) | | 6.6 (5.2-8.4) | |
| | None/missing | 7.6 (7.0-8.3) | | 2.5 (2.4-2.7) | | 5.1 (4.5-5.8) | |
| **Household size** | 1 | 7.0 (6.1-8.0) | <0.001 | 0.05 (0.02-0.12) | <0.001 | 7.0 (6.1-7.9) | 0.004 |
| | 2-4 | 7.9 (7.2-8.6) | | 1.9 (1.9-2.0) | | 5.9 (5.3-6.6) | |
| | 5-7 | 9.5 (8.7-11) | | 4.5 (4.4-4.7) | | 5.0 (4.2-6.0) | |
| | 8-10 | 10 (8.8-11) | | 7.0 (6.3-7.9) | | 3.0 (2.0-4.5) | |
| | 11+ | 15 (9.6-22) | | 9.1 (6.7-13) | | 5.4 (1.9-15) | |
| **Day reported** | Sunday | 8.1 (7.3-8.9) | 0.7 | 2.0 (1.8-2.3) | 0.3 | 6.1 (5.3-6.9) | 0.5 |
| | Monday | 8.6 (7.7-9.6) | | 2.4 (2.2-2.7) | | 6.2 (5.3-7.3) | |
| | Tuesday | 7.8 (7.0-8.8) | | 2.2 (1.9-2.5) | | 5.6 (4.8-6.6) | |
| | Wednesday | 8.5 (7.3-9.9) | | 2.2 (2.0-2.5) | | 6.2 (5.1-7.7) | |
| | Thursday | 8.0 (7.0-9.1) | | 2.0 (1.8-2.4) | | 5.9 (5.0-7.0) | |
| | Friday | 7.1 (6.1-8.3) | | 2.0 (1.6-2.4) | | 5.2 (4.2-6.3) | |
| | Saturday | 8.5 (6.5-11) | | 2.6 (2.2-3.0) | | 5.9 (4.1-8.7) | |
| **Total** | | 8.1 (7.6-8.6) | | 2.2 (2.1-2.3) | | 5.9 (5.4-6.4) | |

Table S2. Mean overall, household, and non-household close contact numbers in Western Cape.

**Close contact time (hours) - KwaZulu-Natal**

| | | Overall | | Household members | | Non-household members | |
|---|---|---|---|---|---|---|---|
| | | Mean (95% CI) | p-value | Mean (95% CI) | p-value | Mean (95% CI) | p-value |
| **Sex** | Male | 56 (52-59) | <0.001 | 40 (37-43) | <0.001 | 16 (14-18) | <0.001 |
| | Female | 82 (78-86) | | 67 (64-71) | | 14 (12-16) | |
| **Age** | 15-19 | 71 (62-81) | 0.96 | 52 (42-63) | 0.6 | 19 (15-25) | 0.2 |
| | 20-29 | 74 (69-79) | | 58 (53-63) | | 16 (14-18) | |
| | 30-39 | 70 (64-77) | | 55 (49-61) | | 15 (13-18) | |
| | 40-49 | 63 (58-69) | | 50 (44-56) | | 13 (11-16) | |
| | 50+ | 74 (68-80) | | 61 (56-66) | | 13 (10-17) | |
| **Residence** | Peri-Urban | 70 (66-75) | 0.04 | 54 (50-59) | 0.01 | 16 (13-18) | 0.8 |
| | Urban | 74 (70-78) | | 60 (56-64) | | 14 (13-15) | |
| | Rural | 60 (52-68) | | 43 (37-51) | | 16 (12-22) | |
| **Monthly household income** | Less than R1000 | 72 (66-78) | <0.001 | 61 (56-67) | <0.001 | 11 (9-13) | <0.001 |
| | R1000 - R2500 | 81 (77-86) | | 69 (65-74) | | 12 (10-13) | |
| | R2500 - R5000 | 53 (48-59) | | 33 (30-37) | | 20 (16-26) | |

| | | | | | | | |
|---|---|---|---|---|---|---|---|
| | R5000 - R10000 | 56 (50-63) | | 30 (25-37) | | 26 (22-31) | |
| | More than R10000 | 73 (62-85) | | 41 (32-53) | | 32 (24-41) | |
| | Unknown/missing | 34 (28-40) | | 23 (16-34) | | 10 (6-18) | |
| **Employment** | Full-time | 64 (58-71) | 0.02 | 45 (40-50) | <0.001 | 19 (15-25) | 0.006 |
| | Part-time | 67 (54-82) | | 51 (38-67) | | 16 (12-21) | |
| | None/missing | 73 (70-77) | | 60 (57-63) | | 14 (12-15) | |
| **Household size** | 1 | 47 (40-54) | <0.001 | 36 (30-43) | <0.001 | 11 (8-15) | 0.06 |
| | 2-4 | 59 (52-67) | | 43 (37-49) | | 16 (11-22) | |
| | 5-7 | 68 (65-73) | | 53 (49-57) | | 15 (13-18) | |
| | 8-10 | 74 (69-79) | | 61 (56-66) | | 13 (11-16) | |
| | 11+ | 87 (80-94) | | 71 (64-78) | | 16 (14-18) | |
| **Day reported** | Sunday | 71 (64-78) | 0.9 | 56 (49-63) | 0.7 | 15 (12-18) | 0.2 |
| | Monday | 70 (62-79) | | 53 (47-61) | | 17 (12-25) | |
| | Tuesday | 70 (64-77) | | 58 (51-65) | | 12 (10-15) | |
| | Wednesday | 73 (66-80) | | 58 (51-66) | | 15 (12-17) | |
| | Thursday | 75 (67-84) | | 58 (51-67) | | 17 (14-21) | |

| | | | | |
|---|---|---|---|---|
| | Friday | 67 (61-74) | 53 (47-60) | 13 (11-16) |
| | Saturday | 73 (66-81) | 59 (52-67) | 14 (12-18) |
| **Total** | | 71 (69-74) | 56 (54-59) | 15 (14-16) |

Table S3. Mean overall, household, and non-household close contact time (hours) in KwaZulu-Natal.

**Close contact time (hours) – Western Cape**

| | | Overall | | Household members | | Non-household members | |
|---|---|---|---|---|---|---|---|
| | | Mean (95% CI) | p-value | Mean (95% CI) | p-value | Mean (95% CI) | p-value |
| **Sex** | Male | 49 (46-53) | <0.001 | 27 (24-30) | <0.001 | 22 (20-25) | <0.001 |
| | Female | 60 (56-64) | | 40 (37-43) | | 20 (18-23) | |
| **Age** | 15-19 | 80 (68-95) | <0.001 | 46 (39-53) | <0.001 | 35 (25-49) | <0.001 |
| | 20-29 | 51 (47-56) | | 28 (25-32) | | 23 (19-27) | |
| | 30-39 | 49 (45-53) | | 31 (28-34) | | 18 (16-21) | |
| | 40-49 | 58 (51-66) | | 38 (32-46) | | 20 (16-25) | |

| | | | | | | | |
|---|---|---|---|---|---|---|---|
| | 50+ | 63 (52-77) | | 44 (35-57) | | 19 (12-31) | |
| **Residence** | Peri-Urban | 55 (52-57) | | 33 (31-35) | | 21 (19-24) | |
| | Urban | NA | | NA | | NA | |
| | Rural | NA | | NA | | NA | |
| **Monthly household income** | Less than R1000 | 58 (48-71) | 0.8 | 37 (30-45) | 0.5 | 22 (16-30) | 0.3 |
| | R1000 - R2500 | 54 (49-60) | | 35 (31-40) | | 19 (16-23) | |
| | R2500 - R5000 | 60 (55-66) | | 34 (31-38) | | 26 (23-30) | |
| | R5000 - R10000 | 41 (35-48) | | 24 (22-28) | | 16 (12-24) | |
| | More than R10000 | 51 (42-62) | | 29 (25-35) | | 21 (14-33) | |
| | Unknown/missing | 57 (51-64) | | 39 (33-46) | | 18 (13-24) | |
| **Employment** | Full-time | 53 (49-58) | 0.2 | 28 (25-31) | <0.001 | 26 (22-29) | 0.002 |
| | Part-time | 52 (46-59) | | 31 (26-36) | | 21 (17-25) | |
| | None/missing | 57 (53-62) | | 40 (36-43) | | 18 (15-21) | |
| **Household size** | 1 | 27 (23-32) | <0.001 | 0.6 (0.3-1.3) | <0.001 | 26 (22-31) | 0.005 |
| | 2-4 | 51 (48-54) | | 29 (28-31) | | 21 (19-24) | |
| | 5-7 | 83 (78-88) | | 66 (62-70) | | 17 (13-22) | |

| | | | | | | | |
|---|---|---|---|---|---|---|---|
| | 8-10 | 124 (109-141) | | 114 (97-134) | 10 (5-21) | | |
| | 11+ | 200 (138-291) | | 172 (112-264) | 28 (7-112) | | |
| **Day reported** | Sunday | 53 (46-60) | 0.8 | 34 (30-52) | 0.5 | 18 (14-23) | 0.7 |
| | Monday | 60 (54-78) | | 34 (30-52) | | 26 (21-29) | |
| | Tuesday | 52 (46-67) | | 32 (28-49) | | 20 (16-27) | |
| | Wednesday | 57 (49-52) | | 32 (28-47) | | 24 (18-22) | |
| | Thursday | 54 (48-66) | | 30 (26-43) | | 23 (19-28) | |
| | Friday | 47 (40-45) | | 27 (23-33) | | 20 (14-19) | |
| | Saturday | 60 (52-53) | | 42 (34-37) | | 18 (14-22) | |
| **Total** | | 55 (52-57) | | 33 (31-35) | | 21 (19-24) | |

Table S4. Mean overall, household, and non-household close contact time (hours) in Western Cape.

**Casual contact time (hours) - KwaZulu-Natal**

| | | Overall | | Household members | | Non-household members | |
|---|---|---|---|---|---|---|---|
| | | Mean (95% CI) | p-value | Mean (95% CI) | p-value | Mean (95% CI) | p-value |
| **Sex** | Male | 171 (158-185) | <0.001 | 86 (81-91) | <0.001 | 85 (73-99) | <0.001 |
| | Female | 203 (191-216) | | 119 (114-125) | | 84 (73-97) | |
| **Age** | 15-19 | 230 (195-270) | 0.02 | 100 (86-115) | 0.6 | 130 (98-172) | 0.005 |
| | 20-29 | 204 (188-221) | | 108 (101-116) | | 95 (80-113) | |
| | 30-39 | 182 (164-202) | | 105 (96-114) | | 77 (61-98) | |
| | 40-49 | 170 (144-200) | | 91 (81-101) | | 79 (56-112) | |
| | 50+ | 186 (170-204) | | 114 (107-122) | | 72 (57-91) | |
| **Residence** | Peri-Urban | 177 (164-190) | 0.6 | 98 (92-104) | <0.001 | 79 (67-92) | 0.06 |
| | Urban | 203 (189-217) | | 117 (111-123) | | 86 (73-100) | |
| | Rural | 186 (154-225) | | 74 (65-85) | | 112 (80-156) | |
| **Monthly household income** | Less than R1000 | 150 (136-166) | 0.002 | 91 (85-99) | 0.2 | 59 (46-74) | <0.001 |
| | R1000 - R2500 | 207 (193-222) | | 126 (119-132) | | 82 (69-97) | |
| | R2500 - R5000 | 191 (170-214) | | 85 (77-94) | | 106 (85-131) | |

| | | | | | | | |
|---|---|---|---|---|---|---|---|
| | R5000 - R10000 | 204 (170-245) | | 97 (83-113) | | 108 (76-152) | |
| | More than R10000 | 222 (175-282) | | 91 (72-113) | | 132 (88-197) | |
| | Unknown/missing | 204 (137-303) | | 69 (54-87) | | 135 (71-256) | |
| **Employment** | Full-time | 188 (165-214) | 0.8 | 87 (79-95) | <0.001 | 101 (80-129) | 0.05 |
| | Part-time | 206 (148-286) | | 82 (66-101) | | 124 (72-213) | |
| | None/missing | 191 (181-201) | | 112 (108-117) | | 78 (69-88) | |
| **Household size** | 1 | 157 (124-198) | 0.2 | 65 (56-77) | <0.001 | 91 (61-136) | 0.8 |
| | 2-4 | 176 (152-204) | | 78 (71-87) | | 98 (76-127) | |
| | 5-7 | 182 (164-201) | | 94 (89-100) | | 87 (71-108) | |
| | 8-10 | 204 (187-224) | | 119 (111-128) | | 85 (68-106) | |
| | 11+ | 207 (191-223) | | 136 (126-146) | | 71 (58-87) | |
| **Day reported** | Sunday | 196 (170-225) | 0.8 | 100 (91-110) | 0.1 | 96 (72-127) | 0.3 |
| | Monday | 177 (157-200) | | 106 (96-118) | | 70 (52-95) | |
| | Tuesday | 191 (165-220) | | 113 (101-126) | | 78 (57-107) | |
| | Wednesday | 194 (171-221) | | 106 (96-117) | | 88 (66-118) | |
| | Thursday | 178 (159-200) | | 106 (95-118) | | 72 (55-95) | |

| | | | | |
|---|---|---|---|---|
| | Friday | 199 (176-226) | 103 (94-113) | 96 (74-125) |
| | Saturday | 199 (176-226) | 108 (98-118) | 91 (70-120) |
| **Total** | | 191 (182-200) | 106 (102-110) | 85 (76-94) |

Table S5. Mean overall, household, and non-household casual contact time (hours) in KwaZulu-Natal.

**Casual contact time (hours) – Western Cape**

| | | Overall | | Household members | | Non-household members | |
|---|---|---|---|---|---|---|---|
| | | Mean (95% CI) | p-value | Mean (95% CI) | p-value | Mean (95% CI) | p-value |
| **Sex** | Male | 142 (125-162) | <0.001 | 38 (34-42) | <0.001 | 104 (88-124) | <0.001 |
| | Female | 161 (146-179) | | 56 (52-62) | | 105 (90-122) | |
| **Age** | 15-19 | 177 (154-204) | 0.4 | 56 (50-64) | 0.006 | 121 (98-148) | 0.9 |
| | 20-29 | 141 (123-162) | | 44 (39-49) | | 98 (81-119) | |
| | 30-39 | 162 (140-188) | | 44 (39-50) | | 119 (97-144) | |
| | 40-49 | 141 (115-173) | | 51 (42-61) | | 90 (67-122) | |

| | | | | | | | |
|---|---|---|---|---|---|---|---|
| | 50+ | 123 (83-182) | | 57 (44-74) | | 66 (34-131) | |
| **Residence** | Peri-Urban | 151 (140-164) | | 47 (44-50) | | 105 (93-117) | |
| | Urban | NA | | NA | | NA | |
| | Rural | NA | | NA | | NA | |
| **Monthly household income** | Less than R1000 | 134 (105-171) | 0.1 | 53 (39-73) | 0.2 | 81 (55-118) | 0.05 |
| | R1000 - R2500 | 160 (137-187) | | 56 (49-64) | | 105 (83-131) | |
| | R2500 - R5000 | 166 (144-192) | | 44 (39-49) | | 122 (101-148) | |
| | R5000 - R10000 | 131 (103-166) | | 36 (32-42) | | 95 (68-132) | |
| | More than R10000 | 133 (103-173) | | 46 (37-58) | | 87 (59-128) | |
| | Unknown/missing | 141 (115-172) | | 46 (39-55) | | 95 (70-129) | |
| **Employment** | Full-time | 159 (138-182) | 0.01 | 37 (33-42) | <0.001 | 122 (102-145) | <0.001 |
| | Part-time | 183 (152-221) | | 43 (35-52) | | 140 (111-178) | |
| | None/missing | 128 (116-141) | | 59 (54-64) | | 69 (59-82) | |
| **Household size** | 1 | 95 (73-123) | <0.001 | 12 (8-17) | <0.001 | 83 (62-112) | 0.4 |
| | 2-4 | 154 (139-170) | | 42 (39-44) | | 113 (98-129) | |
| | 5-7 | 186 (159-217) | | 88 (79-98) | | 98 (73-131) | |

| | | | | | | | |
|---|---|---|---|---|---|---|---|
| | 8-10 | 235 (144-383) | | 133 (109-162) | | 102 (37-279) | |
| | 11+ | 364 (183-722) | | 192 (114-325) | | 171 (35-829) | |
| **Day reported** | Sunday | 150 (123-51) | 0.9 | 49 (42-49) | 0.2 | 100 (75-32) | 0.8 |
| | Monday | 156 (130-55) | | 47 (41-59) | | 110 (85-36) | |
| | Tuesday | 148 (122-50) | | 43 (37-55) | | 105 (80-34) | |
| | Wednesday | 146 (120-50) | | 45 (39-52) | | 100 (75-31) | |
| | Thursday | 137 (108-42) | | 46 (37-33) | | 90 (65-27) | |
| | Friday | 149 (113-36) | | 48 (37-32) | | 101 (69-24) | |
| | Saturday | 175 (141-47) | | 49 (39-34) | | 126 (93-32) | |
| **Total** | | 151 (140-164) | | 47 (44-50) | | 105 (93-117) | |

Table S6. Mean overall, household, and non-household casual contact time (hours) in Western Cape.

#### Details of locations visited and transport used

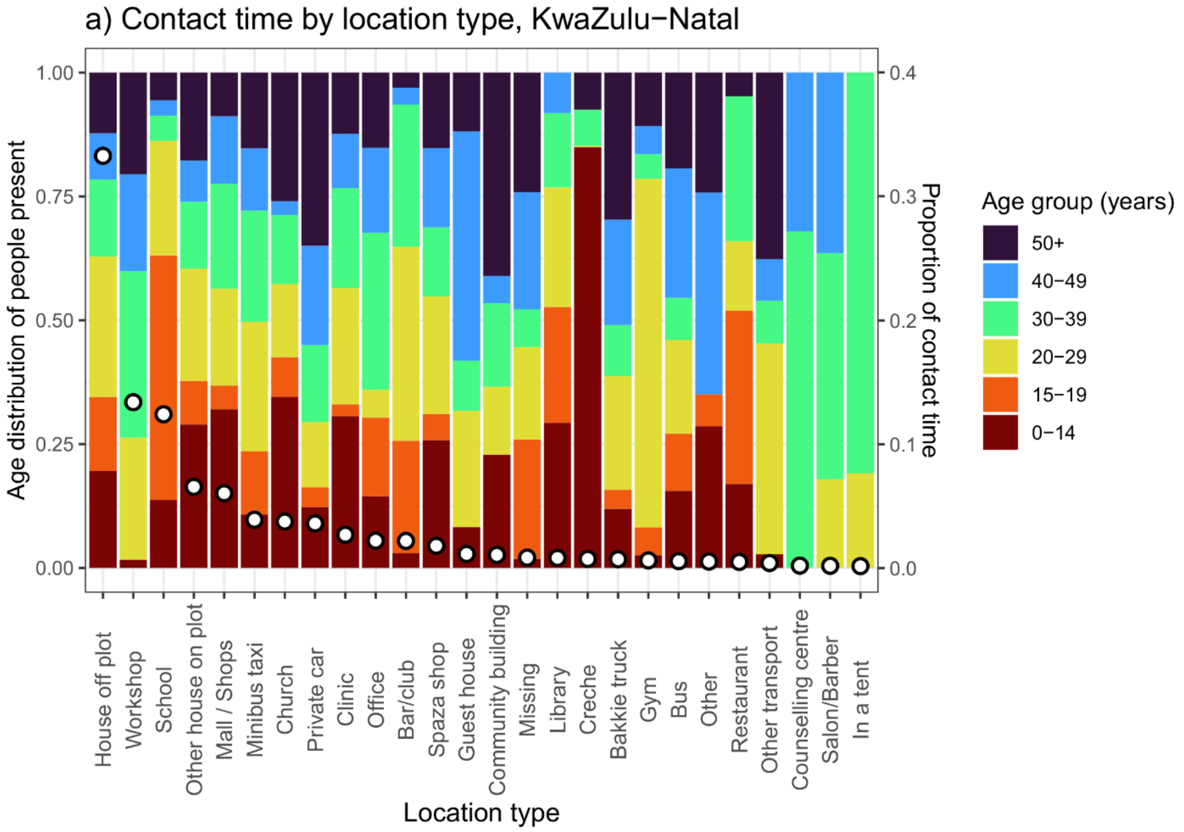

a) Contact time by location type, KwaZulu−Natal
Age distribution of people present
Proportion of contact time
1.00
0.75
0.50
0.25
0.00
0.4
0.3
0.2
0.1
0.0
House off plot
Workshop
School
Other house on plot
Mall / Shops
Minibus taxi
Church
Private car
Clinic
Office
Bar/club
Spaza shop
Guest house
Community building
Missing
Library
Creche
Bakkie truck
Gym
Bus
Other
Restaurant
Other transport
Counselling centre
Salon/Barber
In a tent
Location type
Age group (years)
50+
40−49
30−39
20−29
15−19
0−14


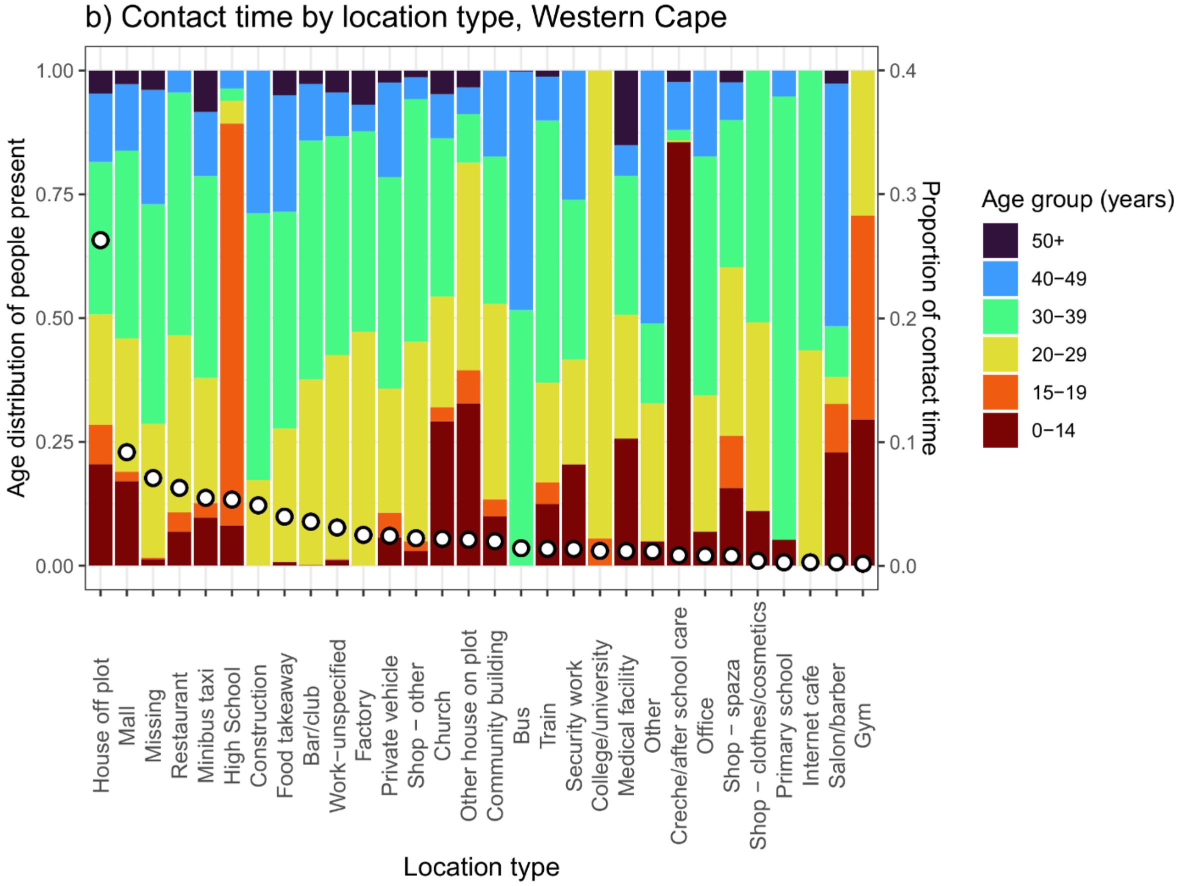

b) Contact time by location type, Western Cape
Age distribution of people present
Proportion of contact time
1.00
0.75
0.50
0.25
0.00
0.4
0.3
0.2
0.1
0.0
House off plot
Mall
Missing
Restaurant
Minibus taxi
High School
Construction
Food takeaway
Bar/club
Work−unspecified
Factory
Private vehicle
Shop − other
Church
Other house on plot
Community building
Bus
Train
Security work
College/university
Medical facility
Other
Creche/after school care
Office
Shop − spaza
Shop − clothes/cosmetics
Primary school
Internet cafe
Salon/barber
Gym
Location type
Age group (years)
50+
40−49
30−39
20−29
15−19
0−14

Figure S5. Age distribution of people present and proportion of contact time by location type, in KwaZulu-Natal and Western Cape. Bars (left axis) show the estimated proportion of people present in a location who are in each age group. Points (right axis) show the estimated proportion of all contact time by adults in the community (excluding people's own homes) that occurs in each type of location.

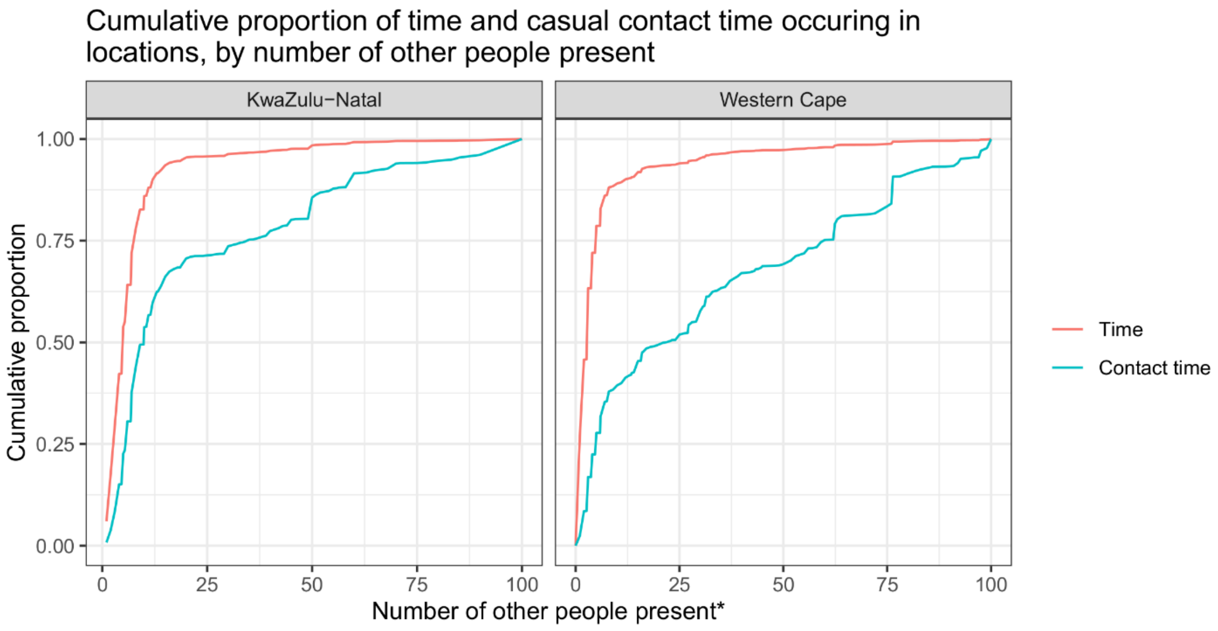


Figure S6. Cumulative proportion of time and casual contact time occurring in locations, by number of other people present, in KwaZulu-Natal and Western Cape. *Number of other people present is capped at 100

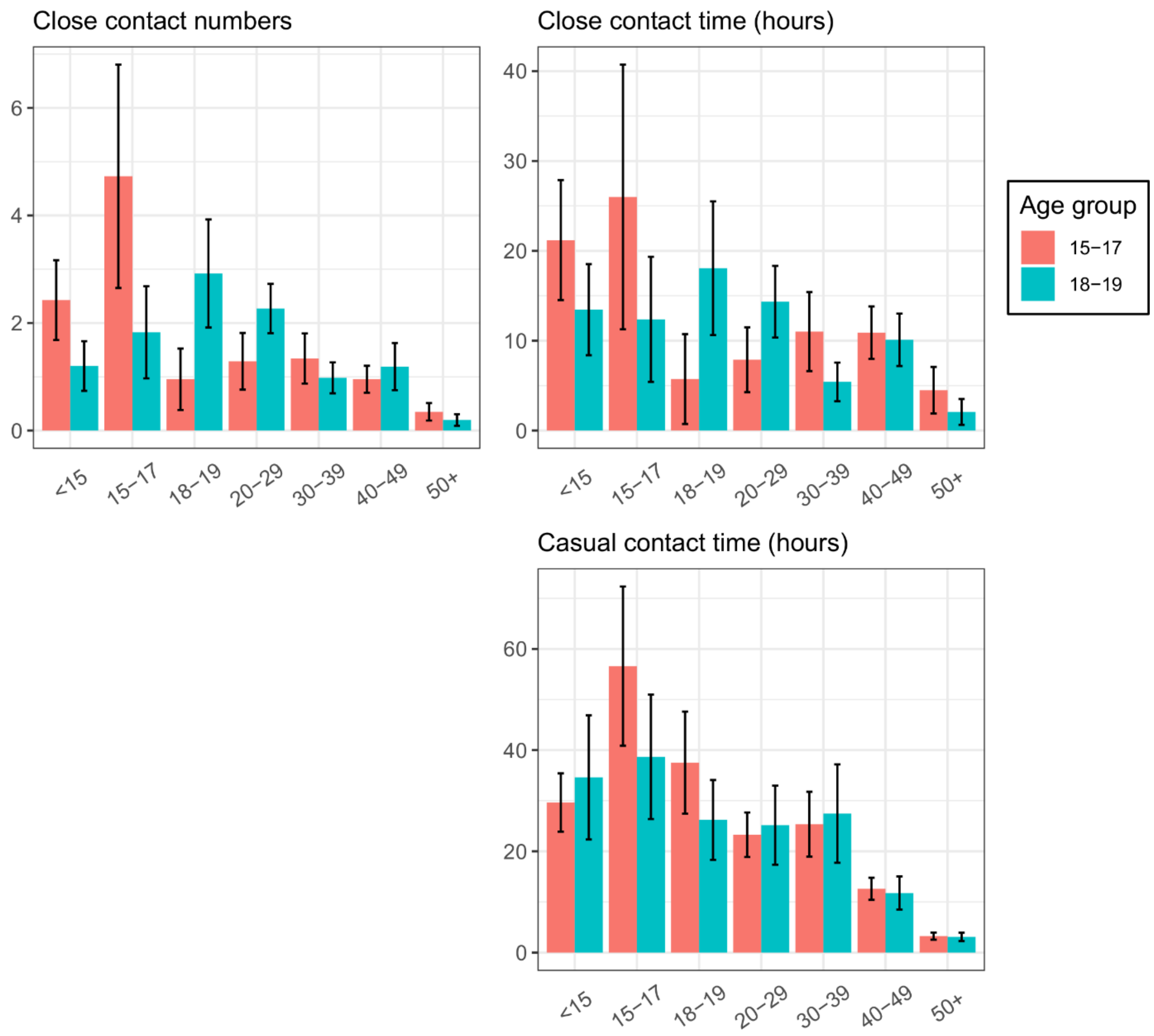


Figure S7. Mean daily close contact numbers, close contact time, and casual contact time by contact age group for respondents aged 15-17 years and 18-19 years in Western Cape. Error bars show 95% confidence intervals.

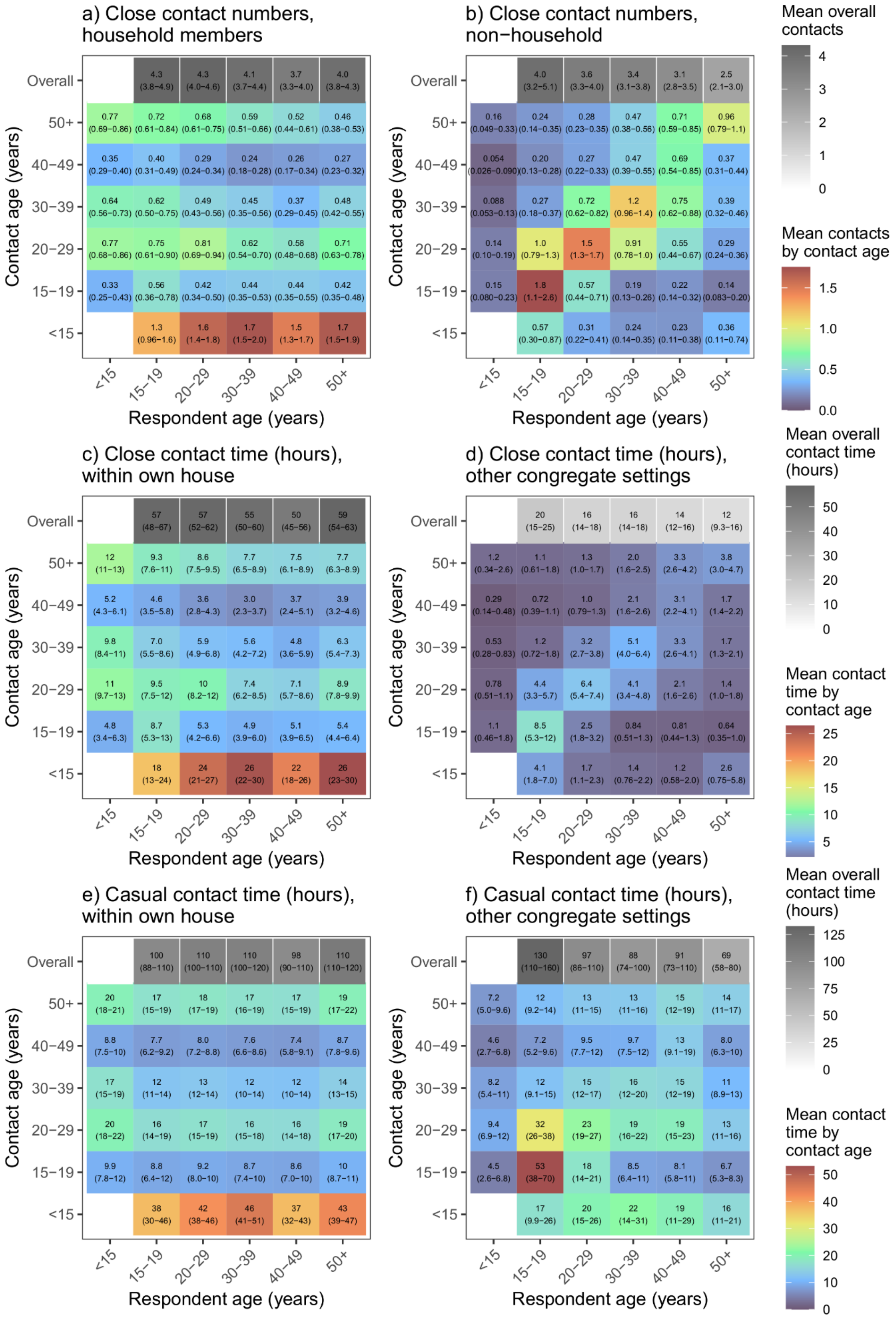

a) Close contact numbers, household members
b) Close contact numbers, non−household
c) Close contact time (hours), within own house
d) Close contact time (hours), other congregate settings
e) Casual contact time (hours), within own house
f) Casual contact time (hours), other congregate settings
Contact age (years)
Respondent age (years)
Overall
50+
40−49
30−39
20−29
15−19
<15
Mean overall contacts
Mean contacts by contact age
Mean overall contact time (hours)
Mean contact time by contact age

Figure S8. Household and non-household close contact numbers, close contact time, and casual contact time age-mixing matrices for KwaZulu-Natal. Numbers shown in graphs a and b are the mean number of contacts respondents in each age group have with contacts in each age group per day. Numbers shown in graphs c-f are the mean number of contact hours respondents in each age group have with contacts in each age group per day. Contact numbers between child 'respondents' and contacts in each age group were estimated from data on contact between adult respondents and child contacts. Ranges shown are bootstrapped 95% plausible ranges.

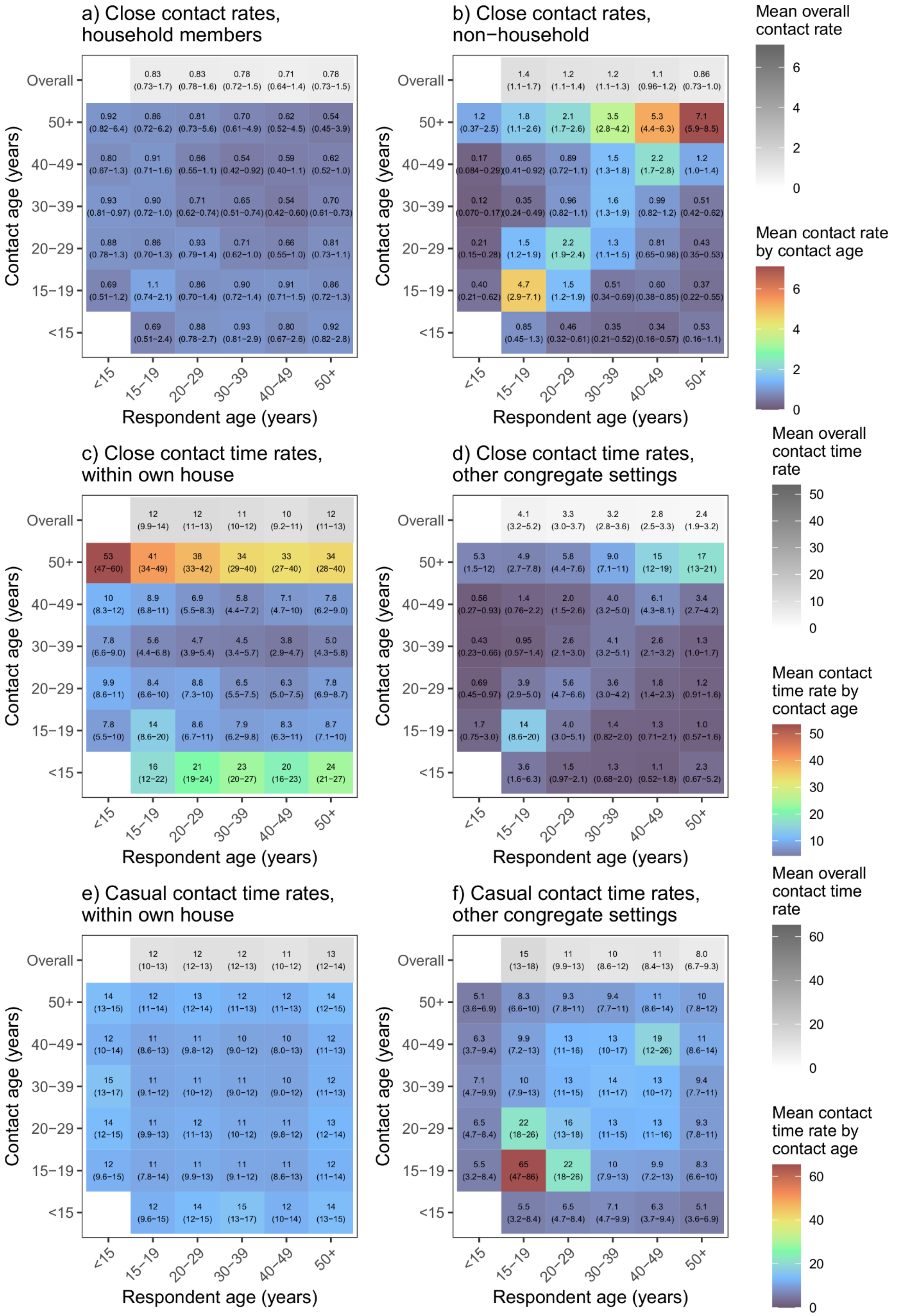
a) Close contact rates, household members
b) Close contact rates, non−household
c) Close contact time rates, within own house
d) Close contact time rates, other congregate settings
e) Casual contact time rates, within own house
f) Casual contact time rates, other congregate settings
Contact age (years)
Respondent age (years)
Overall
50+
40−49
30−39
20−29
15−19
<15
Mean overall contact rate
Mean contact rate by contact age
Mean overall contact time rate
Mean contact time rate by contact age

Figure S9. Household and non-household close contact numbers, close contact time, and casual contact time age-mixing matrices for KwaZulu-Natal. Numbers shown in graphs a and b are the rate of contact between each individual in the population per day, expressed as rates $\times 10^5$. Numbers shown in graphs c-f are the mean minutes of contact between each individual in the population per day, expressed as rates $\times 10^2$. Contact rates between child 'respondents' and contacts in each age group were estimated from data on contact between adult respondents and child contacts. Ranges shown are bootstrapped 95% plausible ranges.

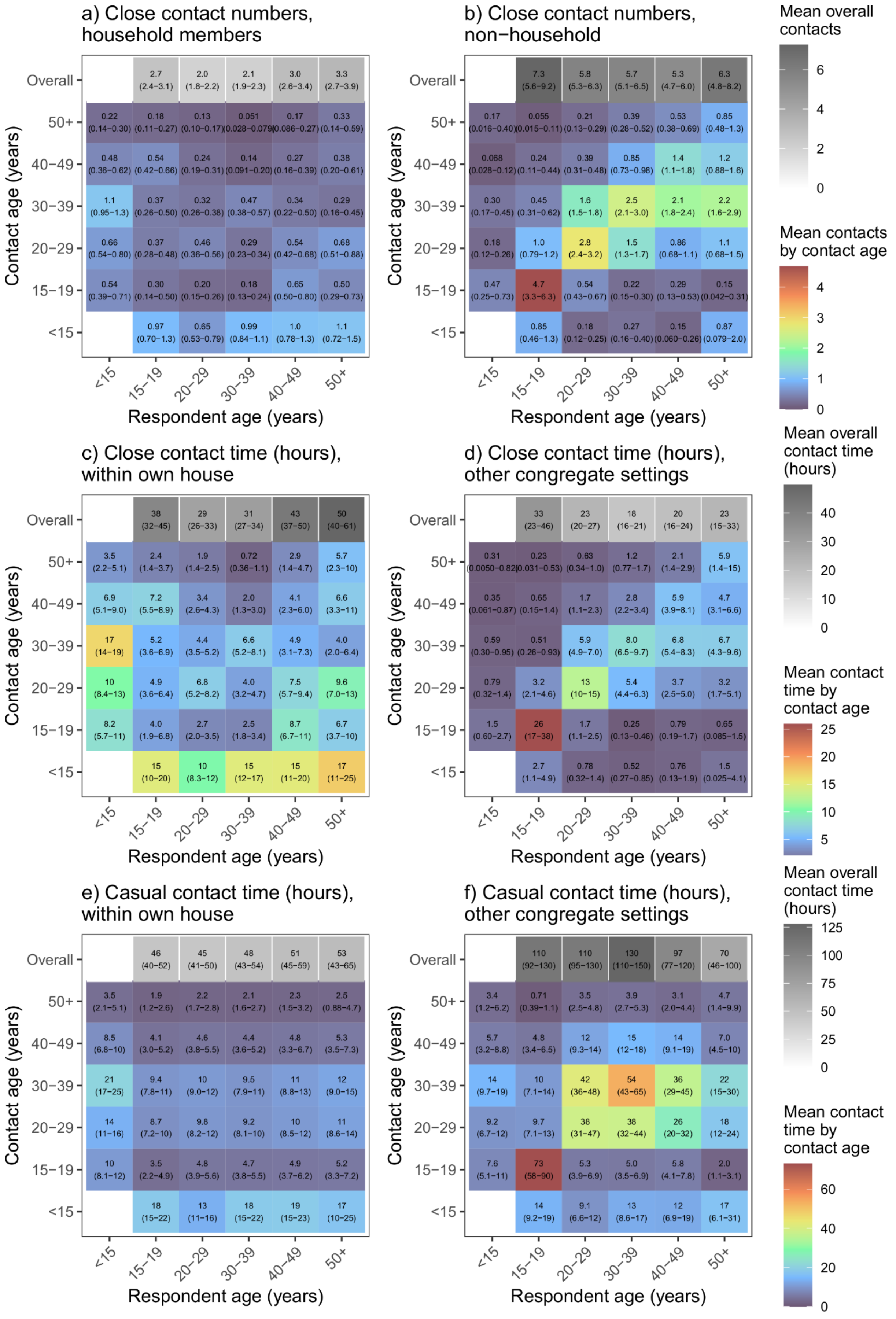
a) Close contact numbers, household members
Overall 2.7 (2.4–3.1) 2.0 (1.8–2.2) 2.1 (1.9–2.3) 3.0 (2.6–3.4) 3.3 (2.7–3.9)
50+ 0.22 (0.14–0.30) 0.18 (0.11–0.27) 0.13 (0.10–0.17) 0.051 (0.028–0.079) 0.17 (0.086–0.27) 0.33 (0.14–0.59)
40−49 0.48 (0.36–0.62) 0.54 (0.42–0.66) 0.24 (0.19–0.31) 0.14 (0.091–0.20) 0.27 (0.16–0.39) 0.38 (0.20–0.61)
30−39 1.1 (0.95–1.3) 0.37 (0.26–0.50) 0.32 (0.26–0.38) 0.47 (0.38–0.57) 0.34 (0.22–0.50) 0.29 (0.16–0.45)
20−29 0.66 (0.54–0.80) 0.37 (0.28–0.48) 0.46 (0.36–0.56) 0.29 (0.23–0.34) 0.54 (0.42–0.68) 0.68 (0.51–0.88)
15−19 0.54 (0.39–0.71) 0.30 (0.14–0.50) 0.20 (0.15–0.26) 0.18 (0.13–0.24) 0.65 (0.50–0.80) 0.50 (0.29–0.73)
<15 0.97 (0.70–1.3) 0.65 (0.53–0.79) 0.99 (0.84–1.1) 1.0 (0.78–1.3) 1.1 (0.72–1.5)
b) Close contact numbers, non−household
Overall 7.3 (5.6–9.2) 5.8 (5.3–6.3) 5.7 (5.1–6.5) 5.3 (4.7–6.0) 6.3 (4.8–8.2)
50+ 0.17 (0.016–0.40) 0.055 (0.015–0.11) 0.21 (0.13–0.29) 0.39 (0.28–0.52) 0.53 (0.38–0.69) 0.85 (0.48–1.3)
40−49 0.068 (0.028–0.12) 0.24 (0.11–0.44) 0.39 (0.31–0.48) 0.85 (0.73–0.98) 1.4 (1.1–1.8) 1.2 (0.88–1.6)
30−39 0.30 (0.17–0.45) 0.45 (0.31–0.62) 1.6 (1.5–1.8) 2.5 (2.1–3.0) 2.1 (1.8–2.4) 2.2 (1.6–2.9)
20−29 0.18 (0.12–0.26) 1.0 (0.79–1.2) 2.8 (2.4–3.2) 1.5 (1.3–1.7) 0.86 (0.68–1.1) 1.1 (0.68–1.5)
15−19 0.47 (0.25–0.73) 4.7 (3.3–6.3) 0.54 (0.43–0.67) 0.22 (0.15–0.30) 0.29 (0.13–0.53) 0.15 (0.042–0.31)
<15 0.85 (0.46–1.3) 0.18 (0.12–0.25) 0.27 (0.16–0.40) 0.15 (0.060–0.26) 0.87 (0.079–2.0)
Mean overall contacts
Mean contacts by contact age
c) Close contact time (hours), within own house
Overall 38 (32–45) 29 (26–33) 31 (27–34) 43 (37–50) 50 (40–61)
50+ 3.5 (2.2–5.1) 2.4 (1.4–3.7) 1.9 (1.4–2.5) 0.72 (0.36–1.1) 2.9 (1.4–4.7) 5.7 (2.3–10)
40−49 6.9 (5.1–9.0) 7.2 (5.5–8.9) 3.4 (2.6–4.3) 2.0 (1.3–3.0) 4.1 (2.3–6.0) 6.6 (3.3–11)
30−39 17 (14–19) 5.2 (3.6–6.9) 4.4 (3.5–5.2) 6.6 (5.2–8.1) 4.9 (3.1–7.3) 4.0 (2.0–6.4)
20−29 10 (8.4–13) 4.9 (3.6–6.4) 6.8 (5.2–8.2) 4.0 (3.2–4.7) 7.5 (5.7–9.4) 9.6 (7.0–13)
15−19 8.2 (5.7–11) 4.0 (1.9–6.8) 2.7 (2.0–3.5) 2.5 (1.8–3.4) 8.7 (6.7–11) 6.7 (3.7–10)
<15 15 (10–20) 10 (8.3–12) 15 (12–17) 15 (11–20) 17 (11–25)
d) Close contact time (hours), other congregate settings
Overall 33 (23–46) 23 (20–27) 18 (16–21) 20 (16–24) 23 (15–33)
50+ 0.31 (0.0050–0.82) 0.23 (0.031–0.53) 0.63 (0.34–1.0) 1.2 (0.77–1.7) 2.1 (1.4–2.9) 5.9 (1.4–15)
40−49 0.35 (0.061–0.87) 0.65 (0.15–1.4) 1.7 (1.1–2.3) 2.8 (2.2–3.4) 5.9 (3.9–8.1) 4.7 (3.1–6.6)
30−39 0.59 (0.30–0.95) 0.51 (0.26–0.93) 5.9 (4.9–7.0) 8.0 (6.5–9.7) 6.8 (5.4–8.3) 6.7 (4.3–9.6)
20−29 0.79 (0.32–1.4) 3.2 (2.1–4.6) 13 (10–15) 5.4 (4.4–6.3) 3.7 (2.5–5.0) 3.2 (1.7–5.1)
15−19 1.5 (0.60–2.7) 26 (17–38) 1.7 (1.1–2.5) 0.25 (0.13–0.46) 0.79 (0.19–1.7) 0.65 (0.085–1.5)
<15 2.7 (1.1–4.9) 0.78 (0.32–1.4) 0.52 (0.27–0.85) 0.76 (0.13–1.9) 1.5 (0.025–4.1)
Mean overall contact time (hours)
Mean contact time by contact age
e) Casual contact time (hours), within own house
Overall 46 (40–52) 45 (41–50) 48 (43–54) 51 (45–59) 53 (43–65)
50+ 3.5 (2.1–5.1) 1.9 (1.2–2.6) 2.2 (1.7–2.8) 2.1 (1.6–2.7) 2.3 (1.5–3.2) 2.5 (0.88–4.7)
40−49 8.5 (6.8–10) 4.1 (3.0–5.2) 4.6 (3.8–5.5) 4.4 (3.6–5.2) 4.8 (3.3–6.7) 5.3 (3.5–7.3)
30−39 21 (17–25) 9.4 (7.8–11) 10 (9.0–12) 9.5 (7.9–11) 11 (8.8–13) 12 (9.0–15)
20−29 14 (11–16) 8.7 (7.2–10) 9.8 (8.2–12) 9.2 (8.1–10) 10 (8.5–12) 11 (8.6–14)
15−19 10 (8.1–12) 3.5 (2.2–4.9) 4.8 (3.9–5.6) 4.7 (3.8–5.5) 4.9 (3.7–6.2) 5.2 (3.3–7.2)
<15 18 (15–22) 13 (11–16) 18 (15–22) 19 (15–23) 17 (10–25)
f) Casual contact time (hours), other congregate settings
Overall 110 (92–130) 110 (95–130) 130 (110–150) 97 (77–120) 70 (46–100)
50+ 3.4 (1.2–6.2) 0.71 (0.39–1.1) 3.5 (2.5–4.8) 3.9 (2.7–5.3) 3.1 (2.0–4.4) 4.7 (1.4–9.9)
40−49 5.7 (3.2–8.8) 4.8 (3.4–6.5) 12 (9.3–14) 15 (12–18) 14 (9.1–19) 7.0 (4.5–10)
30−39 14 (9.7–19) 10 (7.1–14) 42 (36–48) 54 (43–65) 36 (29–45) 22 (15–30)
20−29 9.2 (6.7–12) 9.7 (7.1–13) 38 (31–47) 38 (32–44) 26 (20–32) 18 (12–24)
15−19 7.6 (5.1–11) 73 (58–90) 5.3 (3.9–6.9) 5.0 (3.5–6.9) 5.8 (4.1–7.8) 2.0 (1.1–3.1)
<15 14 (9.2–19) 9.1 (6.6–12) 13 (8.6–17) 12 (6.9–19) 17 (6.1–31)
Mean overall contact time (hours)
Mean contact time by contact age
Contact age (years)
Respondent age (years)
<15 15−19 20−29 30−39 40−49 50+

Figure S10. Household and non-household close contact numbers, close contact time, and casual contact time age-mixing matrices for Western Cape. Numbers shown in graphs a and b are the mean number of contacts respondents in each age group have with contacts in each age group per day. Numbers shown in graphs c-f are the mean number of contact hours respondents in each age group have with contacts in each age group per day. Contact numbers between child 'respondents' and contacts in each age group were estimated from data on contact between adult respondents and child contacts. Ranges shown are bootstrapped 95% plausible ranges.

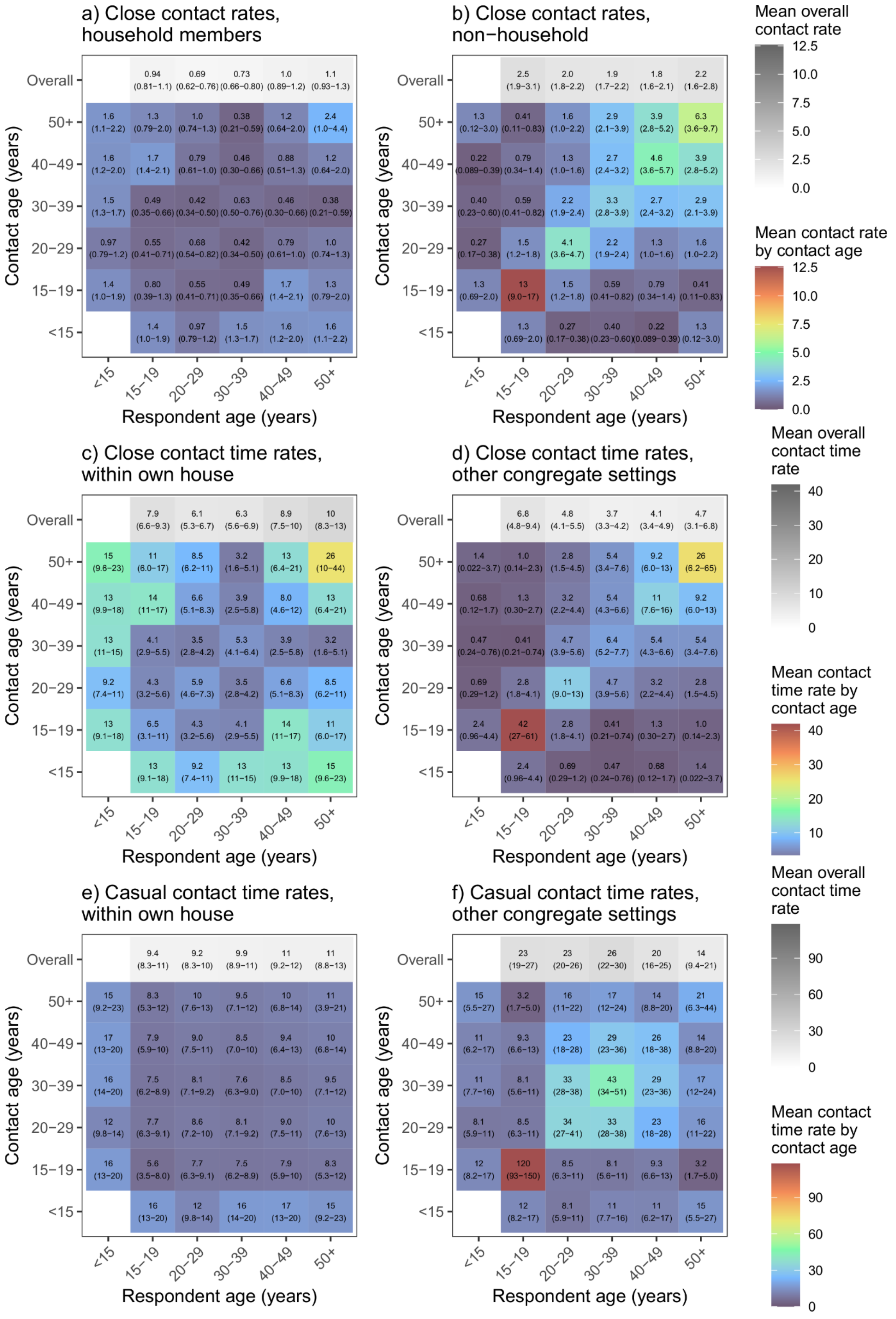
a) Close contact rates, household members
b) Close contact rates, non−household
c) Close contact time rates, within own house
d) Close contact time rates, other congregate settings
e) Casual contact time rates, within own house
f) Casual contact time rates, other congregate settings
Contact age (years)
Respondent age (years)
Overall
50+
40−49
30−39
20−29
15−19
<15
Mean overall contact rate
Mean contact rate by contact age
Mean overall contact time rate
Mean contact time rate by contact age

Figure S11. Household and non-household close contact numbers, close contact time, and casual contact time age-mixing matrices for Western Cape. Numbers shown in graphs a and b are the rate of contact between each individual in the population per day, expressed as rates $\times 10^5$. Numbers shown in graphs c-f are the mean minutes of contact between each individual in the population per day, expressed as rates $\times 10^2$. Contact rates between child 'respondents' and contacts in each age group were estimated from data on contact between adult respondents and child contacts. Ranges shown are bootstrapped 95% plausible ranges.

#### References


1. Funk S. socialmixr: Social Mixing Matrices for Infectious Disease Modelling. The Comprehensive R Archive Network 2018.
2. McCreesh N, Morrow C, Middelkoop K, Wood R, White RG. Estimating age-mixing patterns relevant for the transmission of airborne infections. Epidemics. 2019;28:100339.
3. Ku C-C, MacPherson P, Khundi M, Nzawa R, Feasey HR, Nliwasa M, et al. Estimated durations of asymptomatic, symptomatic, and care-seeking phases of tuberculosis disease. medRxiv. 2021:2021.03.17.21253823.
